# Lifetime air pollution exposure from prenatal to 18 years and cardiovascular health in young adulthood: findings from a UK birth cohort

**DOI:** 10.1101/2025.02.20.25322622

**Authors:** Ana Goncalves Soares, Kate Tilling, Maryam Makanvand, Jules Kerckhoffs, Anna L Hansell, Nicholas J Timpson, Ahmed Elhakeem

**Author notes:** these authors contributed equally (Nicholas J Timpson, Ahmed Elhakeem).

## Abstract

**Aims:** We assessed the association between air pollution from pregnancy (in utero) to 18 years and cardiovascular health markers in early adulthood.

**Methods:** Data from 3,767 individuals from a UK birth cohort were used. We explored the associations between modelled fine particulate matter (PM_2.5_), nitrogen dioxide (NO_2_) and black carbon (BC) across an 18-year period and eight cardiovascular health markers measured at 18 year of age. Long-term exposure to air pollution was assessed by averaging the air pollutants over time and by creating air pollutant trajectories. Linear regressions were used to assess the associations between air pollutants and cardiovascular health markers. Possible sensitive periods of exposure and sex differences in these associations were also explored.

**Results:** Higher average levels of PM_2.5_ and NO_2_ were associated with higher peripheral (pDBP) and central diastolic blood pressure (cDBP); e.g., an interquartile range increase in PM_2.5_ was associated with 0.46 mmHg (95%CI 0.14, 0.78) higher pDBP and 0.50 mmHg (95%CI 0.17, 0.83) higher cDBP. Higher average PM_2.5_ levels were also associated with lower carotid intima-media thickness and higher BC levels were associated with higher heart rate (HR). Latent classes showed the same overall patterns of association, with the trajectory classes with the highest levels of air pollution exposure tending to have higher pDBP, cDBP and HR. There was little evidence of sensitive periods of exposure and sex differences in the associations.

**Conclusions:** Higher lifetime exposure to air pollution up to 18 years was associated with markers of poorer cardiovascular health in early adulthood.

## Introduction

The global disease burden associated with ambient air pollution has considerably increased in the last decades, and the number of disability-adjusted life-years (DALYs) attributable to ambient particulate matter rose by 70% from 1990 to 2019,^1^ including in youths and young adults.^2^ In 2021, particulate matter was the leading contributor to the global disease burden, contributing 8% of total DALYs,^3^ and it was the fourth most important risk factor in relation to the global burden of cardiovascular disease (CVD).^4^ Air pollution has been consistently associated with CVD morbidity and mortality in adulthood^1,5–7^ and this association is now considered causal, supported by evidence of multiple potential mechanisms.^8^ Fewer studies have assessed the impact of air pollution on cardiovascular health in younger ages,^9,10^ and little is known about cardiovascular risk associated with lifetime exposure to air pollution.^11^

Short-(hours to days) and long-term (months to years) exposures to air pollution are associated with higher blood pressure/hypertension,^7^ including in children and adolescents,^9,10^ and associations with intermediate markers of CVD have also been observed. Fine particulate matter (particulate matter ≤ 2.5 µm [PM_2.5_]) has been associated with higher carotid intima-media thickness (CIMT) in adulthood,^12^ though results are less consistent in younger adults.^7,13,14^ Short-term exposure to PM_2.5_ and short– and long-term exposure to nitrogen dioxide (NO_2_) were associated with higher pulse wave velocity (PWV) and augmentation index (AIx) in adults,^15^ including young adults,^16^ but less evidence was observed for other air pollutants.

Long-term exposure to air pollution over years is often assessed by averaging annual mean levels of exposure at someone’s residential address. Although this captures some aspects of cumulative exposure, it does not capture air pollution variability over time and whether this differs for different groups of individuals. Furthermore, exploring different age periods of exposure is important, as previous studies have suggested potential sensitive windows of exposure to air pollution in relation to blood pressure in childhood.^17^ Although previous evidence has shown associations between long-term air pollution and poorer cardiovascular health in adulthood,^1,5–7^ it is not clear how lifetime exposure to air pollution associates with later cardiovascular health.

Some studies have shown possible sex differences in the association between air pollution and CVD. A recent meta-analysis found that the association between PM_2.5_ and ischemic heart disease was stronger in females, whilst the association with stroke was similar in both sexes.^18^ On the other hand, it has been suggested that associations between air pollution and blood pressure might be stronger in males.^7,10,17^ Less evidence for sex differences is found in younger ages,^19^ but studies exploring such differences are scarce.

This study aimed to assess the associations between lifetime exposure to air pollution from pregnancy (prenatal exposure) to 18 years and eight measures of cardiovascular health in young adults. We also explored possible sensitive periods of exposure and potential sex differences in these associations.

## Methods

The Avon Longitudinal Study of Parents and Children (ALSPAC) is a prospective population-based cohort that recruited pregnant women living in the former county of Avon in the United Kingdom who were due to give birth between April 1991 and December 1992.^20,21^ In total, 14,541 pregnancies were initially enrolled, and children, mothers and their partners have been followed up repeatedly ever since. When the oldest children were approximately 7 years of age, an attempt was made to bolster the initial sample with eligible cases who had failed to join the study originally, resulting in a total of 15,447 pregnancies and 15,658 foetuses, from which 14,901 were alive at 1 year of age.^21^ The study website contains details of all the data that is available through a fully searchable data dictionary and a variable search tool (see http://www.bristol.ac.uk/alspac/researchers/our-data for full details of the data from the ALSPAC study).

At average age 17.8 years, 5,217 participants attended a research clinic assessment when a comprehensive range of cardiovascular health indicators were measured. In this study, we included participants who had information on at least one cardiovascular health marker at 18 years, had data on air pollution, and had complete data on confounders. This resulted in the inclusion of 3,767 participants (Supplementary Figure 1). Sample sizes for analyses varied by outcome since not all individuals had all cardiovascular markers measured.

Ethical approval for the study was obtained from the ALSPAC Ethics and Law Committee and the Local Research Ethics Committees. Informed consent for the use of data collected via questionnaires and clinics was obtained from participants following the recommendations of the ALSPAC Ethics and Law Committee at the time. Consent for biological samples has been collected in accordance with the Human Tissue Act (2004). More information on the study ethics is available at http://www.bristol.ac.uk/alspac/researchers/research-ethics/.

### Air pollution exposures

Environmental exposure maps for air pollution were created in the ELAPSE Project (http://www.elapseproject.eu) using land use regression (LUR) models to estimate annual mean PM_2.5_, NO_2_, and black carbon (BC) for 2010.^22^ The models used routine monitoring data from AirBase (v8 dataset)^23^ for PM_2.5_ and NO_2_, and ESCAPE^24^ monitoring data for BC, together with satellite observations, dispersion model estimates, land use information, and traffic data. Kriging was performed on the residual spatial variation from the LUR models and added to the exposure estimates. Values were estimated at 100m resolution.^22^

Participants’ address history records were cleaned and geocoded at the postcode centroid level to 1-m Easting and Northing cartesian coordinates. Detailed information on the geocoding process is available elsewhere.^25^ The exposure to air pollution was estimated at each individual’s address at each year by back-extrapolation using a ratio method with 2010 as the baseline as described previously.^26^ In brief, back-extrapolation used estimated concentrations from the Danish Eulerian Hemispheric Model (DEHM)^27^ applied across Europe at monthly time resolution and 26km x 26km spatial resolution, and the concentration ratio between a year and 2010 from the DEHM model was used to multiply all cohort exposure for that year. The concentration ratios were calculated and applied for each calendar year from pregnancy (between 1990-1992) to age 18 (between 2008-2010). For participants who moved between two years, the exposure was attributed to the mid-point, such that exposure was linked to the previous geocoded address for half of the year and the new address for the other half.

Lifetime exposure to air pollution from pregnancy (prenatal exposure) to 18 years was assessed in two ways: by averaging yearly mean estimates of air pollution, and by creating latent classes (subgroups) using latent class growth analysis (more details described below). The first approach is frequently used in studies assessing long-term exposure to air pollution, and the latter was used to capture the variation in air pollution over time and to identify subgroups of individuals sharing a similar trajectory of exposure.

### Cardiovascular health markers

Eight cardiovascular health markers were used as outcomes: central systolic blood pressure (cSBP), peripheral systolic blood pressure (pSBP), central diastolic blood pressure (cDBP), peripheral diastolic blood pressure (pDBP), heart rate (HR), CIMT, PWV, and AIx. These were measured in a clinical assessment at 18 years by trained fieldworkers using standard protocols.

Brachial (peripheral) blood pressure (mmHg) was measured three times in the right arm, and the average of the second and third readings was used. Central blood pressure (mmHg) and central AIx (%) were measured twice and the average was calculated. HR (bpm) was measured twice, and the average of these measures was used. Baseline (end diastole) CIMT (mm) was measured three times each on the right and left sides and recorded to the nearest 0.01mm; the average of these measures was used. For analysis, CIMT was converted to µm. Carotid to femoral PWV was measured three times and recorded in meters per second (m/s); the average of these measures was calculated.

More details on how each outcome was assessed are presented in supplementary material.

### Covariates

Confounders were determined *a priori* based on a directed acyclic graph and were defined as known or plausible determinants of air pollution (i.e., determinants of where one lives, and therefore of air pollution levels) and cardiovascular health markers. These included maternal age, education and ethnicity, and index of multiple deprivation, all assessed at recruitment. Sex and age at outcome assessment, although not confounders, were included as covariates to reduce residual variance in the outcome. Details on how the covariates were assessed are described in supplementary material.

### Statistical analysis

Average air pollution exposure from pregnancy (perinatal period) to 18 years was calculated by averaging yearly mean estimates. Since not all individuals had complete data for air pollutants at all time points between pregnancy and 18 years, to maximize sample size we included those who had at least 50% of the measures (i.e., at least 10 out of the 19 measures) and at least one measure between ages 0-5, 6-12 and 13-18 and calculated the mean disregarding missing values. For comparison, the average air pollution exposure was also calculated using those with complete data. Pearson correlation between the exposures across ages was calculated. Before analysis, the average levels of air pollutants were rescaled by subtracting the median and dividing by the interquartile range (IQR).

Latent class growth analysis^28,29^ was used to identify subgroups of participants according to air pollution trajectories from pregnancy to 18 years. This model assumes that individuals assigned to the same group (latent class) share the same trajectory of exposure. To select the optimal number of classes, models varying from 2 to 6 classes were fitted for each air pollutant. We used restricted cubic splines with 2 knots to allow for non-linear changes in air pollutants with time.^30^ For each model, a grid of 50 initial values was tested. The same individuals included in the analyses using average levels of air pollution were included in this analysis under the missing at random (MAR) assumption. To select the best model, we considered: i. the quality of adjustment measured by Akaike Information Criterion (AIC) and Bayesian Information Criterion (BIC), with lower values indicating better fit, ii. the relevance of the model and avoidance of small classes (< 5%), and iii. discrimination capacity of the models, based on relative entropy (the closer to one, the better) and average posterior probability (> 0.8).^31^

We described the air pollutants average levels and latent classes according to socioeconomic indicators (i.e., maternal education and area deprivation) to understand the socioeconomic pattern of lifetime air pollution exposure.

Linear regression models adjusted for the covariates defined above were used to assess associations between the average levels of air pollutants from pregnancy to 18 years and the cardiovascular health markers. To assess the associations between air pollution latent classes and the outcomes, weighted linear regressions were used, where each individual was weighted by the posterior probability of belonging to their class, to account for uncertainty of the classes’ classification. Mean differences in the outcomes and their respective 95% confidence intervals (95%CI) were obtained from the regression models.

Potential sensitive windows of exposure were explored by assessing air pollution exposure in different age periods. Mean levels of air pollution between ages 0-5, 6-12 and 13-18 were calculated and the associations between air pollution exposure in each age period and the cardiovascular health markers were assessed using linear regressions adjusted for the covariates defined above. Estimates for the different age ranges were then compared using seemingly unrelated regressions. Given the IQR for each air pollutant at each age period would differ, we performed this analysis using air pollutants in their original scale.

To explore possible sex differences in the associations between air pollution and cardiovascular health markers, an interaction term between each air pollutant average level or latent classes and sex was added to the linear regression models. We also performed sex-stratified analyses.

Association analyses were performed using complete cases, which can produce unbiased results when the chance of being a complete case is independent of the outcome after taking the covariates into consideration.^32^ To explore the potential impact of missing data, we compared characteristics of those included and not included in the analysis due to missing data on air pollutants and covariates.

The analyses were performed in Stata 18.0 (Statacorp, College Station, TX, USA) and R 4.3.1 using the packages *lcmm, splines,* and *lme4*.

### Sensitivity analysis

To explore possible residual confounding in the associations between air pollutants and cardiovascular health markers, we performed a negative control analysis^33^ using birthweight and maternal pre-pregnancy body mass index (BMI) as negative control outcomes. These outcomes have a similar confounding structure to the cardiovascular health markers and are not plausibly influenced by the exposures of interest. Although higher prenatal air pollution exposure has been associated with lower birthweight,^34,35^ postnatal air pollution does not plausibly affect birthweight. Linear regression models adjusted for the same covariates used in the main analysis (except age) were used in the negative control outcome analysis.

Given changes in residential address (spatial variation) explain most of the variability in air pollution levels, we added an interaction term between air pollution and moving status and also stratified the analysis by “movers” (those who moved address at least once between pregnancy and 18 years) and “non-movers” (those who remained in the same address).

## Results

From the 4,935 participants who had data on at least one cardiovascular marker at age 18, 378 did not have data on air pollution and 790 did not have complete data on all confounders, and therefore 3,767 individuals were included in the analysis (Supplementary Figure 1), with sample sizes ranging from 2,960 (for associations with AIx) to 3,574 (for associations with CIMT). The characteristics of the participants included and not included in the analysis due to missing air pollution and/or confounders are described in Table 1. Those included in the analysis were more likely to have lower area deprivation, lower maternal age at birth, higher birthweight and younger age at outcome assessment, and were less likely to have moved address from pregnancy to 18 years than those not included in the analysis. The cardiovascular health markers were similar in those included and not included in the analysis due to missing air pollution and/or confounders, and the average levels of PM_2.5_ and NO_2_ from pregnancy to age 18 were slightly higher in those included in the analysis.

**Table 1.**
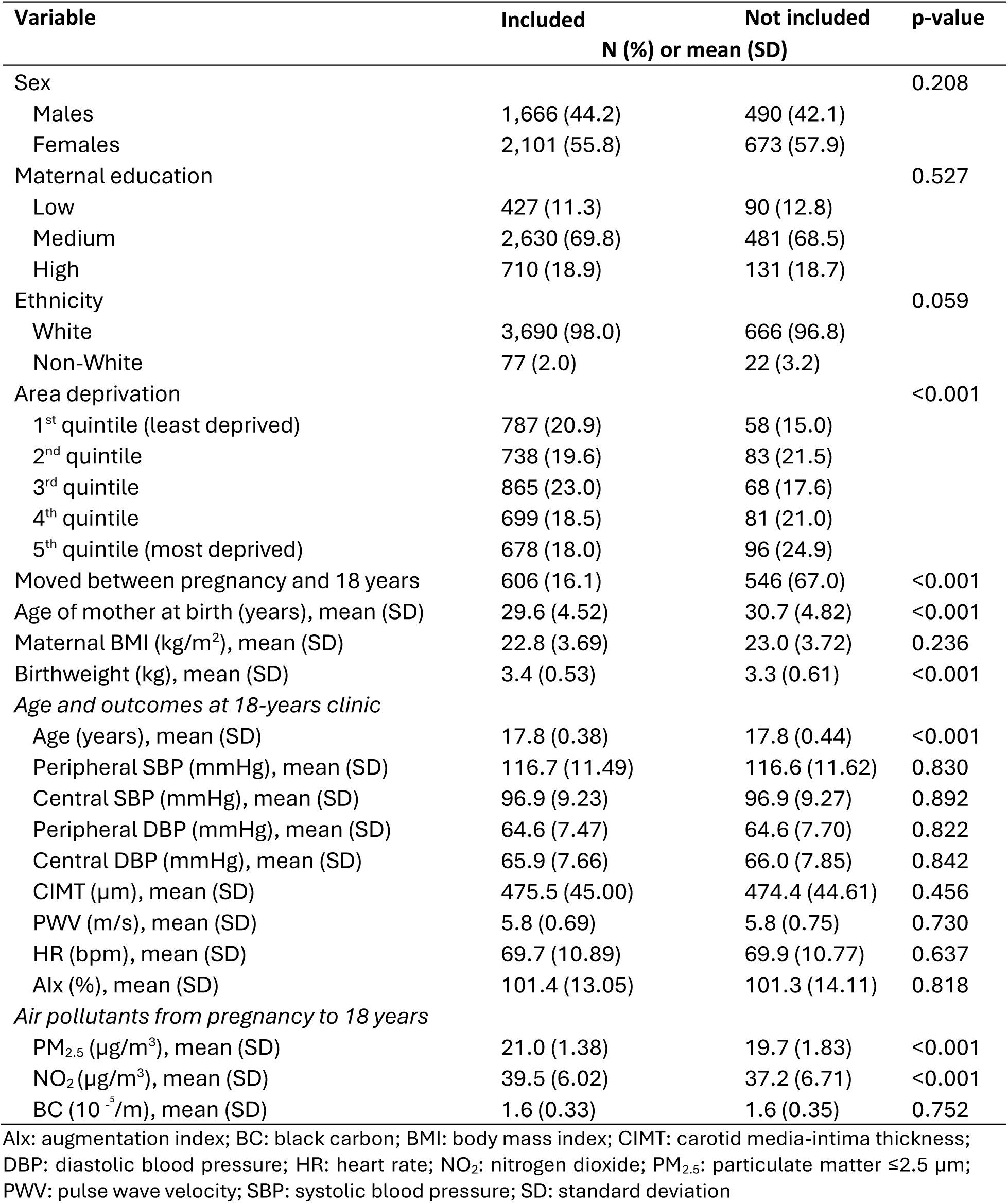
Characteristics of the participants included (n=3,767) and not included in the analysis (n=1,168) due to missing data on air pollution or on one or more of the confounders.

### Lifetime air pollution exposure from pregnancy to 18 years

Most air pollutants had a strong, positive correlation across time, though some exceptions were observed for PM_2.5_ (Supplementary Figure 2). The median PM_2.5_ level from pregnancy (years 1990-1992) to 18 years (years 2008-2010) was 21.1 µg/m^3^ (IQR 1.63), 40.1 µg/m^3^ (IQR 7.96) for NO_2_, and 1.57 x 10^-⁵^/m (IQR 0.55) for BC, and it was similar when only individuals with complete data on air pollution were included (Supplementary Table 1). Lower maternal education and higher area deprivation were associated with higher levels of air pollution, and those who moved address from pregnancy to 18 years had lower levels of air pollution (Supplementary Table 2).

Four latent trajectory classes of PM_2.5_ and six classes of NO_2_ and BC were identified (Figure 1, Supplementary Table 3). For PM_2.5_, the classes had different mean levels but overall followed a similar pattern of decrease from pregnancy to 18 years, and most individuals were classified into the “average-high, decreasing” class (53.5%). For NO_2_, five classes had similar patterns of decrease over time and one class (“high, decreasing fast”, which had 6.8% of the participants) had a faster decrease, especially before 8 years. For BC, four classes had a similar pattern of change over time, with a slight increase followed by a gradual decrease; one class (“low, decreasing”, with 14.7% of the participants) had the lowest level of BC and a constant slow decrease over time; and another class (“high, decreasing fast”, with 7.5% of the participants) had a high level of BC at pregnancy and a fast decrease over time, especially before 10 years. The mean levels of the air pollutants in the latent classes at different ages and across the 18-year period are presented in Supplementary Tables 4-6. Similarly to observed with the average levels, socioeconomic indicators were associated with the air pollution classes such that lower maternal education and higher deprivation were associated with classes with higher levels of air pollution (Supplementary Tables 7-9). Those who moved address often belonged to classes with lower levels of air pollution, or classes with high levels at pregnancy and fast decrease over time (Supplementary Tables 7-9).

**Figure 1.**
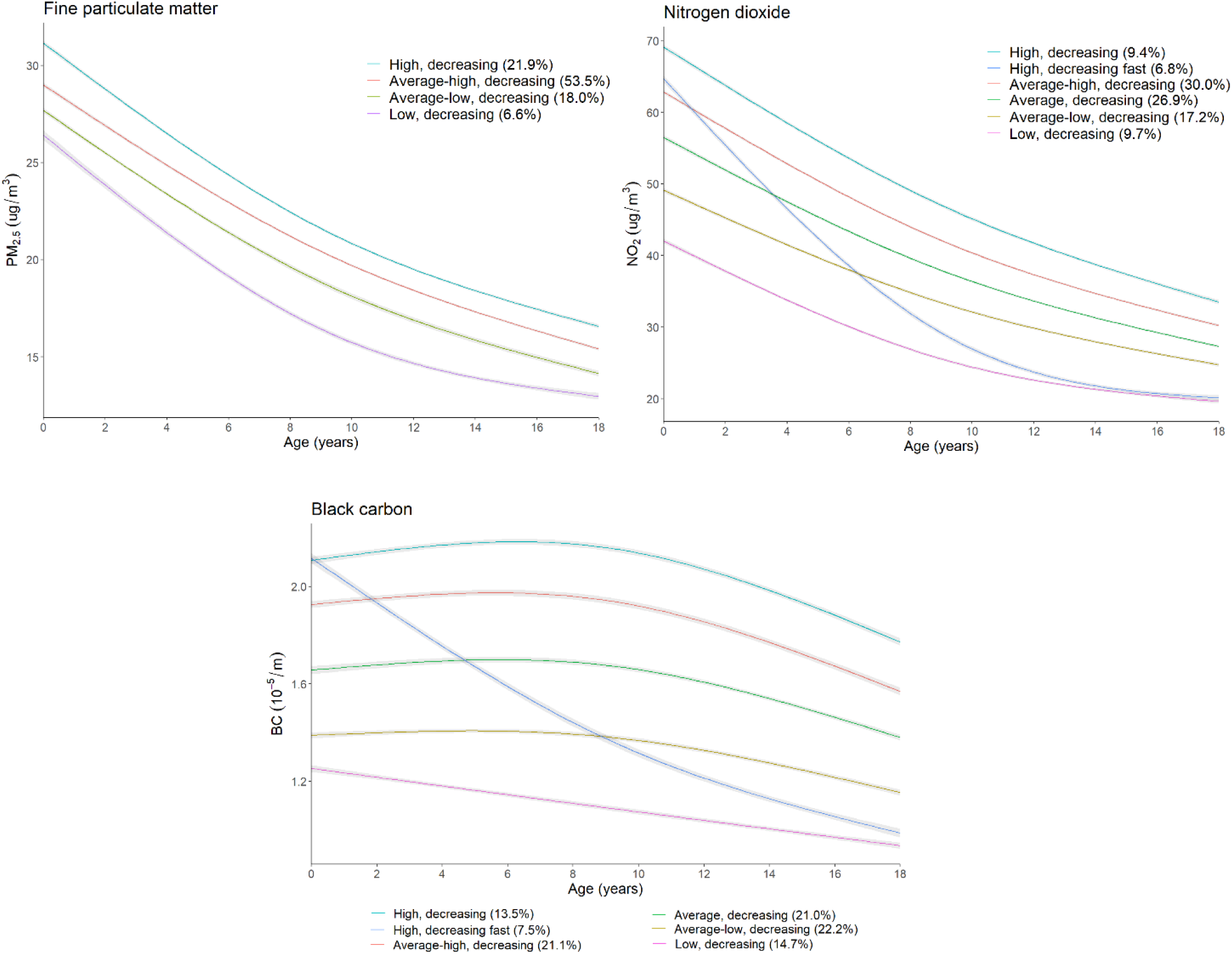
Trajectories of air pollutants from pregnancy to 18 years.

### Associations between lifetime average levels of air pollution and cardiovascular health

Higher lifetime average PM_2.5_ level from pregnancy to 18 years was associated with higher pDBP (mean change in pDBP per IQR increase in lifetime PM_2.5_: 0.46 mmHg, 95% CI 0.14, 0.78) and cDBP (0.50 mmHg, 95%CI 0.17, 0.83), but with lower CIMT (–2.28 µm, 95%CI –4.03, –0.52) (Figure 2, Supplementary Table 10). Higher average PM_2.5_ level was also associated with higher cSBP (0.31 mmHg, 95%CI –0.07, 0.68) and PWV (0.02 m/s, 95%CI –0.01, 0.05), but confidence intervals included the null. Less evidence of association was observed between lifetime average PM_2.5_ levels and the other cardiovascular health markers.

**Figure 2.**
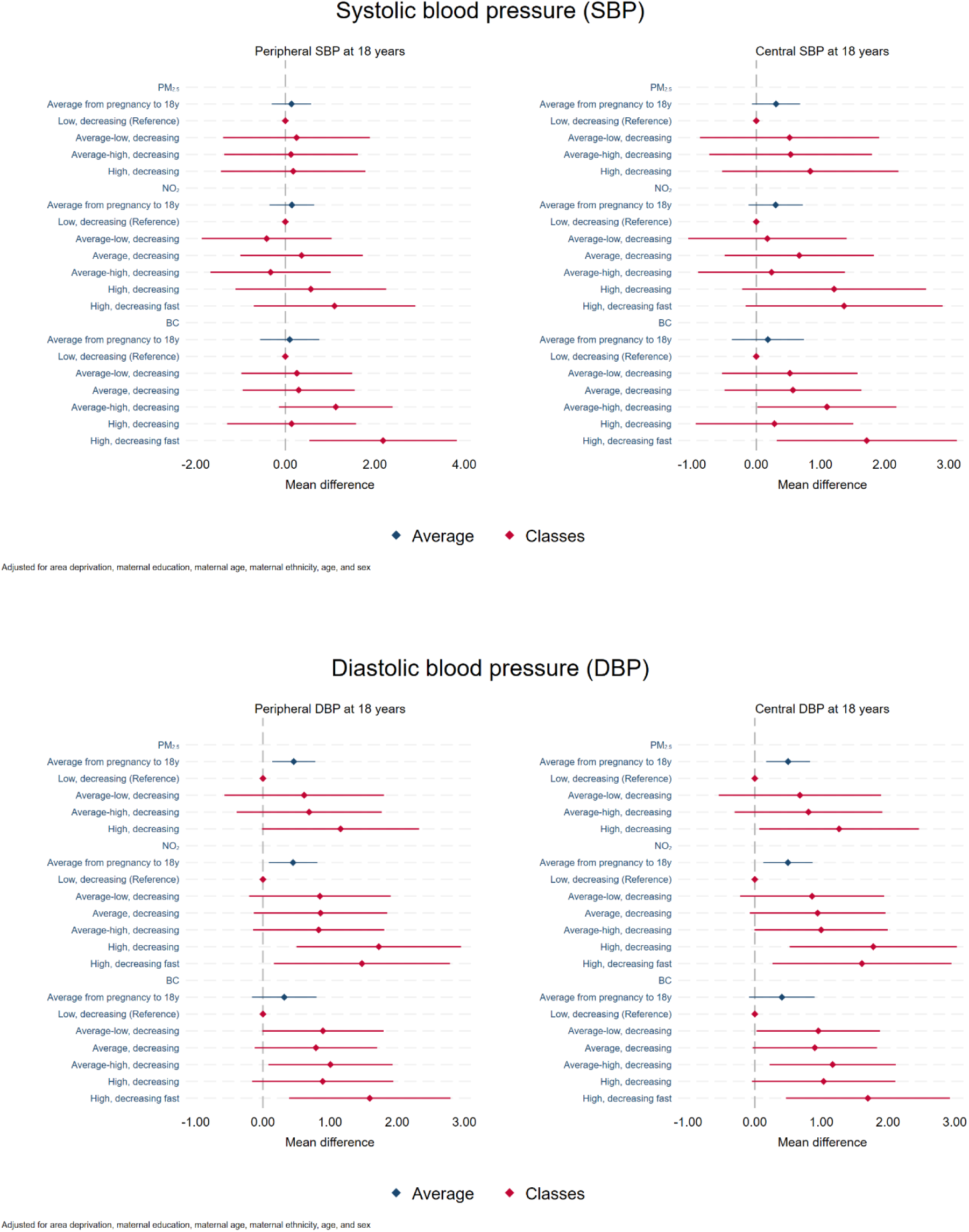
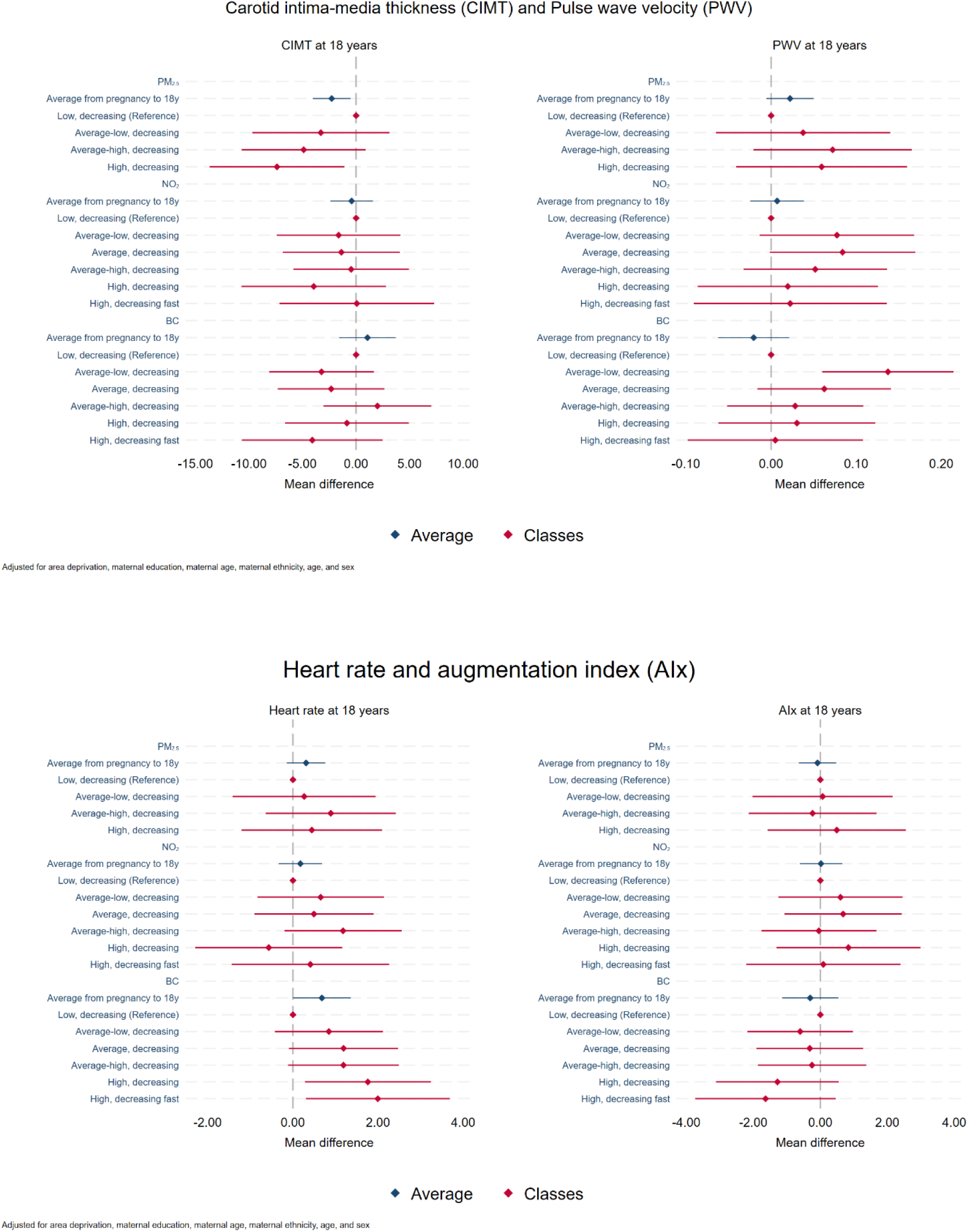
Association between lifetime exposure to air pollution and cardiovascular health markers.

Higher average NO_2_ level from pregnancy to 18 years was associated with higher pDBP (mean change in pDBP per IQR increase in lifetime NO_2_: 0.45 mmHg, 95%CI 0.09, 0.81) and cDBP (0.50 mmHg, 95%CI 0.13, 0.87) (Figure 2, Supplementary Table 11). Although confidence interval included the null, there was also suggestion that higher average NO_2_ level was associated with higher cSBP (0.30 mmHg, 95%CI –0.12, 0.72). There was little evidence to support associations between lifetime average NO_2_ levels and other cardiovascular health markers at age 18.

The association between lifetime average BC and DBP followed the same pattern observed for the other air pollutants, though confidence intervals included the null (mean change per IQR increase in lifetime BC: 0.32 mmHg for pDBP, 95%CI –0.16, 0.80, and 0.41 mmHg for cDBP, 95%CI –0.09, 0.90) (Figure 2, Supplementary Table 12). Higher average BC level from pregnancy to 18 years was associated with higher HR (0.68 bpm, 95%CI 0.00, 1.36) and there was less evidence of associations between lifetime average BC levels and the other cardiovascular health markers.

### Associations between air pollution trajectory classes and cardiovascular health

A similar pattern of association to the ones observed for PM_2.5_ average levels is observed for the latent classes, though some confidence intervals were wide and overlapped the null (Figure 2, Supplementary Table 10). Individuals in the “high, decreasing” PM_2.5_ class (21.9% of the population) had higher cSPB (0.84 mmHg, 95%CI –0.53, 2.21), pDBP (1.16 mmHg, 95%CI –0.01, 2.33) and cDBP (1.26 mmHg, 95%CI 0.07, 2.46), and lower CIMT (–7.39 µm Hg, 95%CI –13.68, –1.10) at age 18 years than those in the “low, decreasing” PM_2.5_ class.

The associations with NO_2_ classes also followed a similar pattern to the ones observed for NO_2_ average levels, such that those in the NO_2_ classes “high, decreasing” (9.4% of the population) and “high, decreasing fast” (6.8% of the population) had higher cSBP, pDBP and cDBP at age 18 than those in the “low, decreasing” NO_2_ class (Figure 2, Supplementary Table 11). E.g., individuals in the “high, decreasing” NO_2_ class had 1.78 mmHg (95%CI 0.52, 3.03) higher cDBP and those in the “high, decreasing fast” class had 1.61 mmHg (95%CI 0.26, 2.95) higher cDBP compared to those in the “low, decreasing” class, although there was little evidence to support differences in the associations across the different NO_2_ latent classes (p = 0.095).

BC latent classes also showed a similar pattern of association with HR than seen for BC average levels, although there was little evidence to support differences in the associations across the different classes (p = 0.159). E.g., individuals classified into the BC classes “high-decreasing” (which had 13.5% of the population), “high, decreasing fast” (7.5% of the population) and those in the “average-high, decreasing” class (21.1% of the population) had higher HR than those in the “low, decreasing” BC class, by 1.99 bpm (95%CI 0.30, 3.68), 1.76 bpm (95%CI 0.28, 3.24) and 1.19 bpm (95%CI –0.12, 2.49), respectively (Figure 2, Supplementary Table 12). Although some confidence intervals included the null, there is also possibility of adverse associations with pSBP, cSBP, pDBP and cDBP.

### Sensitive periods of exposure to air pollution

Supplementary Figure 3 shows the correlation between the exposures at different age ranges and the associations between air pollution exposure at different age periods and cardiovascular health markers at 18 years are presented in Supplementary Figure 4 and Supplementary Table 13. Associations between air pollution and markers of cardiovascular health were overall similar across ages, and there was little evidence of sensitive periods of exposure.

### Sex differences

Most associations between air pollutants and cardiovascular health markers were similar in males and females (Supplementary Tables 14-16). The only exceptions were for the associations of higher average BC levels from pregnancy to 18 years with higher pDBP and cDBP at 18 years, which were evident only in males (0.78mmHg, 95%CI 0.06, 1.51, and 0.92mmHg, 95%CI 0.18, 1.67, respectively) but not in females (–0.07mmHg, 95%CI –0.71, 0.58, and – 0.02mmHg, 95%CI –0.68, 0.64, respectively) (p-values for sex interaction: p= 0.030 and p=0.024, respectively).

### Comparing movers and non-movers

Although some associations between air pollutants and cardiovascular health markers were only observed in those who moved addresses at least once between pregnancy and 18 years, there was little evidence to support differences in the associations between those who moved and did not move address (Supplementary Tables 17-19).

### Negative control outcome analyses

When we assessed the association between lifetime exposure to air pollutants and maternal pre-pregnancy BMI and birthweight, little evidence of association was observed (Supplementary Table 20), suggesting little evidence of residual confounding.

## Discussion

We assessed the association between lifetime exposure to air pollution from pregnancy (in utero) to 18 years and eight cardiovascular health markers in early adulthood. We found that higher lifetime average levels of PM_2.5_, NO_2_ and BC were associated with higher pDBP and cDBP, higher PM_2.5_ average levels were also associated with lower CIMT, and higher BC average levels were associated with higher HR. Similar patterns of association were also observed in the air pollution trajectory classes with a higher exposure over the 18-year period. There was little evidence to support sensitive periods of exposure and sex differences in the associations between lifetime air pollution exposure and cardiovascular health markers in early adulthood.

The associations observed between lifetime air pollution exposure from pregnancy to age 18 with poorer cardiovascular health are in line with previous findings exploring long-term air pollution and cardiovascular and metabolic health in younger ages and adulthood.^7,9,10,12–16^ The only exception was the association between PM_2.5_ and lower CIMT, which was observed in the opposite direction than previous findings. In adulthood PM_2.5_ has been associated with higher CIMT,^12^ and this has also been observed in children (mean age 11.9 years),^36^ but the association in adolescence and early adulthood seems to be inconsistent. A previous study assessing average monthly estimates of air pollution from birth to 10-11 years also showed negative, though imprecise, associations between PM_2.5_ and CIMT at 21-22 years (β per standard deviation increase in PM_2.5_ –1.62 μm, 95%CI –18.38, 15.14) and yearly changes in CIMT between ages 10– 11 and 21–22 years (–0.40 μm/year, 95%CI – 1.75, 0.96).^14^ On the other hand, another study in young adults (mean age 21.1 years) found that PM_2.5_ was associated with higher CIMT (% change per IQR increase in PM_2.5_ 0.46%, 95%CI: 0.02, 0.90).^13^ A study using quantile regression to assess the association between air pollution from birth up to age 14 and CIMT in adolescents (mean age 16.3 years) found little evidence that PM_2.5_ was associated with CIMT, though higher levels of PM_2.5_ absorbance (a marker for black carbon) at birth was associated with higher CIMT only at the lowest decile of the CIMT distribution.^37^ Additional studies are needed to understand the association between air pollution and CIMT at younger ages. Whilst CIMT is an important biomarker of subclinical atherosclerosis and well-established marker for future adverse CVD outcomes in adults, subtle changes in CIMT in young individuals may represent physiological adaptations as opposed to subclinical atherosclerosis.^38^

Both pre– and postnatal exposure to PM_2.5_ have been associated with higher SBP and DBP in children and adolescents,^10^ but little is known about the cumulative exposure over the life course. In our study, lifetime trajectories of air pollution from pregnancy to 18 years using groups identified via latent class growth analysis showed similar direction of associations than those observed with average annual mean levels of air pollution. Despite being underpowered to show differences in the associations across different latent classes of air pollution, consistent patterns of association and associations in specific classes were observed. Latent class growth analysis allows the identification of sub-groups of individuals who follow a similar trajectory of exposure to air pollution and how the exposure changes over the period in these groups. Although this approach been previously used for air pollution in relation to kidney function,^39^ latent trajectory models have been underexplored when assessing the association between lifetime exposure to air pollution and health outcomes.

Even though estimates were mostly imprecise, our study suggests associations between BC, a combustion-related constituent of PM_2.5_, and adverse cardiovascular health markers in early adulthood. Whilst the association between PM_2.5_ and cardiovascular health has been more widely studied,^5,6,8,40^ the association of PM_2.5_ components has been less investigated.^41,42^ BC has been associated with higher SBP and DBP,^42,43^ including in children and adolescents,^44^ with most studies being cross-sectional. We found that lifetime BC levels might be associated with higher SBP and DBP and is also associated with higher HR in early adulthood.

We found little evidence of sensitive periods in the associations between lifetime air pollution exposure and cardiovascular health in early adulthood. Studies exploring prenatal air pollution suggest that higher exposure during the second and third trimesters of pregnancy seem to be particularly important in relation to offspring’s blood pressure.^17,45^ We only located one study that investigated lifetime exposure to air pollution in relation to cardiometabolic diseases. This examined PM_2.5_ exposure into three stages (<20, 20–40 and >40 years) in adults (mean age 63.4 years) and found that exposure to PM_2.5_ after 40 years was associated with higher risk of metabolic syndrome, hypertension, diabetes and cardiovascular disease.^11^ However, this study did not have information on residential address history and lifetime air pollution estimates were linked to the residential address in adulthood only. Lifetime exposure to air pollution and potential sensitive periods of exposure have been explored in relation to epigenetic ageing^46^ and respiratory health,^47,48^ suggesting that different health outcomes might have different sensitive periods of exposure to air pollution. Further studies exploring lifetime exposure to air pollution, and potential sensitive periods of exposure, in relation to cardiovascular health are needed to fully understand the potential impacts of prolonged exposure to air pollution.

Although previous studies have suggested sex differences in the association between air pollution and cardiovascular health,^7,10,17,18^ we found little evidence to support such differences in the association between air pollution from pregnancy to 18 years and cardiovascular health in early adulthood. The only exception was a stronger association between higher levels of BC and both higher pDBP and cDBP in males compared to females. Most previous studies have performed sex-specific analyses instead of exploring potential effect modification, which can lead to misinterpretation of results, but some studies exploring effect modification have found mixed results.^17,49^ Additional work is needed to elucidate possible sex-specific associations in relation to air pollution and cardiovascular health. If such differences indeed exist, this might help target interventions.

There was little evidence to suggest effect modification by moving status, despite some associations observed were stronger or only evident in those who moved addresses at least once between pregnancy and 18 years. This is expected given spatial variation explains great part of the variability in air pollution levels. In our study, those who moved address were less likely to have been included in the analysis, and were also more likely to have higher socioeconomic status and lower levels of air pollution. We also found little evidence of residual confounding by socioeconomic status in our negative control analysis.

### Strengths and limitations

Our study contributes to the currently very limited evidence on the association between air pollution and cardiovascular health in young adults that considers lifetime exposure to air pollution. We assessed long-term air pollution over an 18-year period from pregnancy (in utero) and explored its association with a range of cardiovascular health indicators in early adulthood. We used air pollution levels linked to residential address history, which reduces misclassification in exposure assignments. Besides assessing average annual mean levels of air pollution we also assessed air pollution trajectories, exploring patters of change in air pollution over time. We explored potential sensitive periods of exposure to air pollution and sex differences in the association between air pollution and cardiovascular health. Additionally, we used negative control analysis to assess residual confounding by socioeconomic status in the associations between air pollution and cardiovascular health.

This study also has several limitations which should be acknowledged. ALSPAC has considerable attrition, with those from more deprived backgrounds and who moved addresses being less likely to participate in the 18-year follow-up. Although the cardiovascular outcomes were similar in those included and not included in the analysis due to missing air pollution and/or confounders data, it is not possible to assess differences in the outcomes in relation to those who did not attend the 18-year follow-up. Therefore, it is possible that associations observed here are biased and likely underestimated. We used back-extrapolation to estimate concentrations of air pollution in different years, which can introduce exposure misclassification, which if non-differential, would likely result in bias towards the null. We did not assess potential interactions between air pollutants, which is likely to occur; this also increases the complexity of the analyses and interpretation of results. We used latent class growth analysis, which is a data-driven approach, and the latent classes identified here might not replicate in other studies. Furthermore, some classes were small, which meant we had limited statistical power to identify differences in the associations between the classes. We used negative control outcomes to assess potential residual confounding, but these analyses should be interpreted with some caution. Although it is implausible that later air pollution exposure would influence earlier outcomes such as maternal BMI and birthweight, it might be difficult to fully disentangle air pollution exposure at different ages given their high correlations. Lastly, despite the number of exposures and outcomes assessed in this study, we did not account for multiple testing. Instead, we interpreted our results based on the patterns of associations and on the magnitude and precision of the estimates.

### Conclusions

This study showed that higher air pollution exposure from pregnancy (in utero) to 18 years is associated with poorer cardiovascular health in early adulthood. These finding are consistent with the extensive body of evidence supporting associations between air pollution and cardiovascular outcomes in later life and raise the potential that exposure to higher levels of air pollution from early life might have long-lasting negative impacts on cardiovascular health.

## Data Availability

Researchers can apply to use ALSPAC data, including the variables under investigation in this study. Data access information is provided here: http://www.bristol.ac.uk/alspac/researchers/access/

## Acknowledgments

We are extremely grateful to all the families who took part in this study, the midwives for their help in recruiting them, and the whole ALSPAC team, which includes interviewers, computer and laboratory technicians, clerical workers, research scientists, volunteers, managers, receptionists and nurses.

## Funding

This project received funding from the European Union’s Horizon 2020 research and innovation programme (874739 LongITools and 874724 Equal-Life). AGS, KT, MM, NTJ and AE work in a Unit that is funded by the UK Medical Research Council (MC_UU_00011/1&6) and the University of Bristol. The UK Medical Research Council and Wellcome (Grant ref: 217065/Z/19/Z), and the University of Bristol provide core support for ALSPAC. A comprehensive list of grants funding is available on the ALSPAC website (http://www.bristol.ac.uk/alspac/external/documents/grant-acknowledgements.pdf). AH acknowledges funding from the National Institute for Health and Care Research (NIHR) Leicester Biomedical Research Centre (BRC), additionally from the NIHR Health Protection Research Unit (HPRU) in Environmental Exposures and Health, a partnership between the UK Health Security Agency (UKHSA), the Health and Safety Executive (HSE), and the University of Leicester (Grant number: NIHR200901) and from a British Heart Foundation (BHF) Research Excellence Award (RE/24/130031). The views expressed are those of the author(s) and not necessarily those of the European Union’s Horizon 2020, UK Medical Research Council, Wellcome, NIHR, Department of Health and Social Care, UKHSA, HSE, or BHF.

## Notes

### Competing Interest Statement

The authors have declared no competing interest.

### Funding Statement

This project received funding from the European Unions Horizon 2020 research and innovation programme (874739 LongITools and 874724 Equal-Life). AGS, KT, MM, NTJ and AE work in a Unit that is funded by the UK Medical Research Council (MC_UU_00011/1&6) and the University of Bristol. The UK Medical Research Council and Wellcome (Grant ref: 217065/Z/19/Z), and the University of Bristol provide core support for ALSPAC. A comprehensive list of grants funding is available on the ALSPAC website (http://www.bristol.ac.uk/alspac/external/documents/grant-acknowledgements.pdf). AH acknowledges funding from the National Institute for Health and Care Research (NIHR) Leicester Biomedical Research Centre (BRC), additionally from the NIHR Health Protection Research Unit (HPRU) in Environmental Exposures and Health, a partnership between the UK Health Security Agency (UKHSA), the Health and Safety Executive (HSE), and the University of Leicester (Grant number: NIHR200901) and from a British Heart Foundation (BHF) Research Excellence Award (RE/24/130031). The views expressed are those of the author(s) and not necessarily those of the European Unions Horizon 2020, UK Medical Research Council, Wellcome, NIHR, Department of Health and Social Care, UKHSA, HSE, or BHF.

### Author Declarations

Ethical approval for the study was obtained from the ALSPAC Ethics and Law Committee and the Local Research Ethics Committees.

