## Supplementary material for "Lifetime air pollution exposure from prenatal to 18 years and cardiovascular health in young adulthood: findings from a UK birth cohort"

**Supplementary methods**

*Cardiovascular health markers assessment*

Carotid intima-media thickness (CIMT) was measured using B-mode ultrasound from ZONARE z.oneUltra convertible ultrasound machines. Both left and right carotid arteries were imaged three times 1-2 cm proximal to the carotid bifurcation. Ten second cine-loops were recorded and analysed offline by trained practitioners. CIMT was recorded to the nearest 0.01 mm, and the average of the right and left side measures was used.

For brachial (peripheral) blood pressure and heart rate, individuals wore appropriate loose sleeves on the upper-arm and were asked to sit in a chair and rest each arm on a table. The circumference of one upper arm was measured in cm. The appropriate sized blood pressure cuff was fitted to the person's upper arm. Brachial blood pressure was measured after resting in a seated position for 5 minutes, using an Omron 705 IT semiautomatic oscillometric recorder (Omron Electronic Components Europe BV). Blood pressure and heart rate were measured three times in the right arm, and the average of the second and third readings was used.

Central blood pressure and central augmentation index (AIx) were measured using radial artery tonometry with a SphygmoCor Px Pulse Wave Analysis System (Atcor Medical). Recordings were calibrated using the brachial blood pressure (measured with the Omron 705 IT in the same session) according to manufacturer's instructions. Central blood pressure (mmHg) and central AIx (%) were measured twice and the average was calculated.

Carotid to femoral pulse wave velocity (PWV) was assessed using Vicorder portable physiologic vascular testing equipment. PWV velocity was calculated from measurements of pulse transit time and distance travelled by the pulse between carotid to femoral. For PWV, a cuff was placed over the right carotid artery in the participant's neck, while another was located over the femoral artery in their upper right thigh. The distance between the participant's suprasternal notch and the top of the thigh cuff was measured, as was the distance between their suprasternal notch and the bottom of the neck cuff on the right side. PWV and transit time were automatically computed. PWV was measured three times and recorded in meters per second (m/s); the average of these measures was used.

*Covariates*

Maternal age, education and ethnicity were assessed at recruitment. Maternal age at delivery was calculated from the mother's date of birth (obtained at enrolment) and the date of delivery. Maternal level of education was based on the highest ongoing or completed education. It was classified as low, medium and high according to the International Standard Classification of Education 97/2011 (ISCED-97/2011). High education corresponded to short cycle tertiary, Bachelor, Masters, Doctoral or equivalent (ISCED2011: 5-8, ISCED-97: 5-6); medium education corresponded to upper secondary, or post-secondary non-tertiary (ISCED-2011: 3-4, ISCED-97: 3-4); and low education corresponded to no education, early childhood, pre-primary, primary, lower secondary or second stage of basic education (ISCED-2011: 0-2, ISCED-97: 0-2). Maternal ethnicity was self-reported at enrolment and recoded into White and non-White.

Index of multiple deprivation (IMD) was based on data from the Ministry of Housing, Communities & Local Government (UK Government) from 2015 and linked to geocoded addresses at recruitment. It combined weighted information from seven domains to produce an overall relative measure of deprivation. The domains and their weights were the following: income deprivation (22.5%), employment deprivation (22.5%), education, skills and training deprivation (13.5%), health deprivation and disability (13.5%), crime (9.3%), barriers to housing and services (9.3%), and living environment deprivation (9.3%). IMD was categorised in quintiles, with the first corresponding to the least deprived and the last to the most deprived.

Sex was obtained from the birth notification at enrolment. Age at outcome assessment (complete years) was calculated from date of birth and date of attendance at the research clinic.

**Supplementary Tables and Figures**

**Supplementary Table 1**. Descriptive statistics of air pollution from pregnancy to 18 years.

**Supplementary Table 2.** Association between socioeconomic indicators and air pollutants averages from pregnancy to 18 years.

**Supplementary Table 3**. Model fit parameters for group-based trajectory models of air pollution from pregnancy to 18 years.

**Supplementary Table 4**. Mean levels of fine particulate matter ($PM_{2.5}$) (µg/$m^3$) in each age by latent classes.

**Supplementary Table 5**. Mean levels of nitrogen dioxide ($NO_2$) (µg/$m^3$) in each age by latent classes.

**Supplementary Table 6**. Mean levels of black carbon ($10^{-5}$/m) in each age by latent classes.

**Supplementary Table 7**. Fine particulate matter ($PM_{2.5}$) classes distribution by socioeconomic indicators.

**Supplementary Table 8**. Nitrogen dioxide ($NO_2$) classes distribution by socioeconomic indicators.

**Supplementary Table 9**. Black carbon (BC) classes distribution by socioeconomic indicators.

**Supplementary Table 10.** Association between fine particulate matter ($PM_{2.5}$) from pregnancy to 18 years with cardiovascular health indicators at age 18 years.

**Supplementary Table 11.** Association between nitrogen dioxide ($NO_2$) from pregnancy to 18 years with cardiovascular health indicators at age 18 years.

**Supplementary Table 12.** Association between black carbon (BC) from pregnancy to 18 years with cardiovascular health indicators at age 18 years.

**Supplementary Table 13.** Association between air pollution at different age ranges and cardiovascular health markers at age 18 years.

**Supplementary Table 14**. Association between fine particulate matter ($PM_{2.5}$) and cardiovascular health markers at 18 years in males and females.

**Supplementary Table 15.** Association between nitrogen dioxide ($NO_2$) and cardiovascular health markers at 18 years in males and females.

**Supplementary Table 16.** Association between black carbon (BC) and cardiovascular health markers at 18 years in males and females.

**Supplementary Table 17**. Association between fine particulate matter ($PM_{2.5}$) and cardiovascular health markers at 18 years in non-movers and movers.

**Supplementary Table 18.** Association between nitrogen dioxide ($NO_2$) and cardiovascular health markers at 18 years in non-movers and movers.

**Supplementary Table 19.** Association between black carbon (BC) and cardiovascular health markers at 18 years in non-movers and movers.

**Supplementary Table 20.** Association between air pollutants from pregnancy to 18 years with birthweight and maternal pre-pregnancy body mass index (BMI).

**Supplementary Figure 1.** Flow chart of participants.

**Supplementary Figure 2.** Correlation between air pollutants across ages from pregnancy (age 0) to 18 years.

**Supplementary Figure 3.** Correlation between air pollutants across age ranges.

**Supplementary Figure 4.** Association between air pollution at different age ranges and cardiovascular health markers at age 18 years.

**Supplementary Table 1**. Descriptive statistics of air pollution from pregnancy to 18 years.

| Pollutant/ age | N | Mean | SD | Median | p25 | p75 | IQR |
|---|---|---|---|---|---|---|---|
| ***$PM_{2.5}$ (μg/m³)*** | | | | | | | |
| 0 | 3,762 | 29.75 | 2.53 | 29.26 | 28.10 | 31.81 | 3.71 |
| 1 | 3,760 | 28.89 | 1.83 | 29.07 | 28.03 | 30.03 | 2.00 |
| 2 | 3,760 | 25.94 | 2.99 | 25.03 | 23.88 | 28.63 | 4.75 |
| 3 | 3,758 | 24.04 | 1.58 | 24.25 | 23.26 | 24.91 | 1.65 |
| 4 | 3,754 | 26.14 | 2.41 | 26.40 | 24.42 | 27.91 | 3.49 |
| 5 | 3,754 | 26.15 | 2.11 | 26.25 | 25.20 | 27.54 | 2.34 |
| 6 | 3,754 | 21.75 | 3.24 | 20.52 | 19.57 | 25.02 | 5.45 |
| 7 | 3,751 | 19.22 | 1.46 | 19.49 | 18.57 | 20.15 | 1.58 |
| 8 | 3,755 | 18.24 | 1.49 | 18.40 | 17.54 | 19.23 | 1.69 |
| 9 | 3,759 | 19.01 | 1.74 | 19.18 | 18.03 | 20.28 | 2.25 |
| 10 | 3,755 | 19.32 | 1.54 | 19.51 | 18.71 | 20.28 | 1.57 |
| 11 | 3,755 | 21.30 | 2.41 | 21.74 | 19.51 | 23.31 | 3.80 |
| 12 | 3,756 | 19.69 | 2.67 | 18.90 | 17.99 | 22.21 | 4.22 |
| 13 | 3,758 | 17.88 | 1.43 | 18.24 | 17.26 | 18.75 | 1.49 |
| 14 | 3,760 | 18.32 | 1.54 | 18.62 | 17.64 | 19.27 | 1.63 |
| 15 | 3,756 | 18.26 | 1.51 | 18.57 | 17.58 | 19.20 | 1.62 |
| 16 | 3,755 | 16.08 | 1.98 | 15.69 | 14.85 | 17.68 | 2.83 |
| 17 | 3,753 | 14.08 | 1.31 | 14.12 | 13.39 | 15.01 | 1.62 |
| 18 | 3,754 | 14.04 | 1.20 | 14.20 | 13.49 | 14.80 | 1.31 |
| Average | 3,767 | 20.95 | 1.38 | 21.11 | 20.26 | 21.89 | 1.63 |
| Average# | 3,717 | 20.96 | 1.37 | 21.12 | 20.27 | 21.89 | 1.63 |
| ***$NO_2$ (μg/m³)*** | | | | | | | |
| 0 | 3,766 | 58.44 | 9.07 | 59.36 | 52.99 | 64.45 | 11.45 |
| 1 | 3,760 | 55.86 | 8.78 | 56.85 | 50.68 | 61.71 | 11.03 |
| 2 | 3,760 | 49.69 | 8.82 | 50.05 | 44.15 | 55.54 | 11.39 |
| 3 | 3,754 | 47.10 | 7.65 | 48.30 | 42.72 | 52.28 | 9.56 |
| 4 | 3,754 | 50.74 | 8.84 | 51.61 | 45.58 | 56.91 | 11.32 |
| 5 | 3,751 | 50.65 | 8.71 | 51.81 | 45.77 | 56.65 | 10.88 |
| 6 | 3,752 | 42.61 | 8.97 | 42.29 | 36.83 | 48.53 | 11.70 |
| 7 | 3,750 | 38.22 | 6.62 | 39.25 | 34.44 | 42.96 | 8.52 |
| 8 | 3,754 | 35.31 | 6.53 | 36.15 | 31.37 | 39.73 | 8.35 |
| 9 | 3,757 | 37.81 | 7.51 | 38.41 | 32.93 | 43.40 | 10.47 |
| 10 | 3,754 | 36.66 | 7.04 | 37.28 | 32.28 | 41.40 | 9.12 |
| 11 | 3,754 | 36.29 | 6.70 | 37.18 | 32.28 | 41.04 | 8.76 |
| 12 | 3,756 | 33.83 | 6.58 | 34.34 | 29.76 | 38.25 | 8.48 |
| 13 | 3,758 | 31.60 | 5.76 | 32.40 | 28.30 | 35.81 | 7.51 |
| 14 | 3,760 | 30.99 | 5.68 | 31.80 | 27.71 | 35.11 | 7.40 |
| 15 | 3,756 | 30.71 | 5.64 | 31.55 | 27.46 | 34.85 | 7.39 |
| 16 | 3,755 | 29.65 | 5.54 | 30.47 | 26.28 | 33.61 | 7.33 |
| 17 | 3,753 | 26.58 | 5.27 | 27.12 | 23.23 | 30.15 | 6.92 |
| 18 | 3,754 | 26.82 | 5.18 | 27.40 | 23.60 | 30.46 | 6.86 |
| Average | 3,767 | 39.45 | 6.02 | 40.15 | 35.78 | 43.74 | 7.96 |
| Average# | 3,715 | 39.45 | 6.00 | 40.15 | 35.79 | 43.74 | 7.95 |

| | | | | | | | |
|---|---|---|---|---|---|---|---|
| ***BC ($10^{-5}$/m)*** | | | | | | | |
| 0 | 3,766 | 1.77 | 0.39 | 1.80 | 1.45 | 2.06 | 0.61 |
| 1 | 3,760 | 1.78 | 0.38 | 1.80 | 1.48 | 2.10 | 0.62 |
| 2 | 3,760 | 1.62 | 0.38 | 1.62 | 1.31 | 1.88 | 0.57 |
| 3 | 3,758 | 1.52 | 0.33 | 1.54 | 1.26 | 1.81 | 0.55 |
| 4 | 3,755 | 1.76 | 0.43 | 1.76 | 1.44 | 2.08 | 0.64 |
| 5 | 3,754 | 1.85 | 0.42 | 1.87 | 1.51 | 2.21 | 0.70 |
| 6 | 3,754 | 1.56 | 0.41 | 1.54 | 1.23 | 1.82 | 0.58 |
| 7 | 3,751 | 1.37 | 0.31 | 1.37 | 1.11 | 1.64 | 0.53 |
| 8 | 3,755 | 1.44 | 0.34 | 1.44 | 1.17 | 1.71 | 0.55 |
| 9 | 3,759 | 1.71 | 0.43 | 1.69 | 1.38 | 2.02 | 0.65 |
| 10 | 3,755 | 1.71 | 0.41 | 1.69 | 1.38 | 2.03 | 0.65 |
| 11 | 3,755 | 1.82 | 0.45 | 1.79 | 1.46 | 2.14 | 0.68 |
| 12 | 3,756 | 1.63 | 0.44 | 1.60 | 1.28 | 1.91 | 0.63 |
| 13 | 3,758 | 1.49 | 0.35 | 1.49 | 1.21 | 1.80 | 0.59 |
| 14 | 3,760 | 1.55 | 0.38 | 1.54 | 1.25 | 1.86 | 0.61 |
| 15 | 3,756 | 1.46 | 0.35 | 1.44 | 1.18 | 1.74 | 0.57 |
| 16 | 3,755 | 1.34 | 0.32 | 1.33 | 1.08 | 1.61 | 0.53 |
| 17 | 3,753 | 1.24 | 0.30 | 1.23 | 0.99 | 1.49 | 0.50 |
| 18 | 3,754 | 1.36 | 0.34 | 1.34 | 1.08 | 1.61 | 0.53 |
| Average | 3,767 | 1.58 | 0.33 | 1.57 | 1.31 | 1.86 | 0.55 |
| Average[#] | 3,722 | 1.58 | 0.33 | 1.57 | 1.31 | 1.87 | 0.55 |

[#] complete cases

BC: black carbon; IQR: interquartile range; $NO_2$: nitrogen dioxide; p25: 25th percentile, p75: 75th percentile, $PM_{2.5}$: fine particulate matter, SD: standard deviation

**Supplementary Table 2.** Association between socioeconomic indicators and air pollutants averages from pregnancy to 18 years.

| Socioeconomic indicators | Difference in mean (95% CI) | | |
|---|---|---|---|
| | **$PM_{2.5}$** | **$NO_2$** | **BC** |
| Maternal education | p<0.001 | p<0.001 | p=0.001 |
| Low | Reference | Reference | Reference |
| Medium | -0.28 (-0.42, -0.14) | -1.09 (-1.70, -0.47) | -0.05 (-0.09, -0.02) |
| High | -0.67 (-0.84, -0.51) | -0.98 (-1.70, -0.26) | -0.05 (-0.09, -0.01) |
| Area deprivation | p<0.001 | p<0.001 | p<0.001 |
| 1$^{st}$ Q (least deprived) | Reference | Reference | Reference |
| 2$^{nd}$ quintile | -0.19 (-0.33, -0.05) | -1.00 (-1.59, -0.42) | 0.01 (-0.02, 0.04) |
| 3$^{rd}$ quintile | 0.10 (-0.03, 0.23) | 1.37 (0.81, 1.93) | 0.14 (0.11, 0.17) |
| 4$^{th}$ quintile | 0.42 (0.28, 0.56) | 3.46 (2.87, 4.05) | 0.27 (0.23, 0.30) |
| 5$^{th}$ Q (most deprived) | 0.54 (0.40, 0.68) | 3.03 (2.43, 3.62) | 0.26 (0.23, 0.29) |
| Moving status from pregnancy to 18 years | p<0.001 | p<0.001 | p<0.001 |
| Moved address | -1.55 (-1.66, -1.44) | -4.82 (-5.32, -4.32) | -0.26 (-0.29, -0.23) |

BC: black carbon; $NO_2$: nitrogen dioxide; $PM_{2.5}$: fine particulate matter; Q: quintile

**Supplementary Table 3.** Model fit parameters for group-based trajectory models of air pollution from pregnancy to 18 years.

| Air pollutant | Classes | BIC | AIC | APP | Entropy | Class 1 (%) | Class 2 (%) | Class 3 (%) | Class 4 (%) | Class 5 (%) | Class 6 (%) |
|---|---|---|---|---|---|---|---|---|---|---|---|
| $PM_{2.5}$ | 1 | 337164.5 | 337139.6 | NA | 1.000 | 100.00 | NA | NA | NA | NA | NA |
| | 2 | 327482.0 | 327432.2 | 0.960 | 0.894 | 23.97 | 76.03 | NA | NA | NA | NA |
| | 3 | 324651.2 | 324576.4 | 0.922 | 0.817 | 11.87 | 33.98 | 54.15 | NA | NA | NA |
| | **4** | **323854.4** | **323754.7** | **0.908** | **0.846** | **7.51** | **34.11** | **53.04** | **5.34** | **NA** | **NA** |
| | 5 | 323145.9 | 323021.2 | 0.879 | 0.807 | 5.23 | 52.24 | 4.91 | 20.92 | 16.70 | NA |
| | 6 | 322959.6 | 322809.9 | 0.859 | 0.781 | 3.34 | 27.48 | 12.95 | 46.08 | 4.99 | 5.15 |
| $NO_2$ | 1 | 490896.5 | 490871.5 | NA | 1.000 | 100.00 | NA | NA | NA | NA | NA |
| | 2 | 456193.6 | 456143.7 | 0.987 | 0.956 | 60.76 | 39.24 | NA | NA | NA | NA |
| | 3 | 443395.4 | 443320.6 | 0.976 | 0.946 | 41.17 | 42.74 | 16.09 | NA | NA | NA |
| | 4 | 437908.9 | 437809.2 | 0.961 | 0.929 | 29.57 | 12.82 | 20.28 | 37.32 | NA | NA |
| | 5 | 432431.3 | 432306.6 | 0.963 | 0.941 | 11.55 | 25.22 | 36.24 | 19.88 | 7.11 | NA |
| | **6** | **429934.1** | **429784.5** | **0.951** | **0.926** | **30.00** | **26.86** | **9.69** | **17.23** | **9.45** | **6.77** |
| BC | 1 | 72506.27 | 72481.33 | NA | 1.000 | 100.00 | NA | NA | NA | NA | NA |
| | 2 | 30689.42 | 30639.55 | 0.989 | 0.963 | 51.69 | 48.31 | NA | NA | NA | NA |
| | 3 | 19442.2 | 19367.39 | 0.974 | 0.946 | 33.71 | 34.54 | 31.75 | NA | NA | NA |
| | 4 | 15313.33 | 15213.59 | 0.960 | 0.931 | 26.92 | 27.32 | 28.01 | 17.76 | NA | NA |
| | 5 | 11434.29 | 11309.61 | 0.959 | 0.940 | 7.43 | 24.63 | 23.17 | 27.48 | 17.28 | NA |
| | **6** | **9603.894** | **9454.277** | **0.940** | **0.917** | **22.25** | **21.05** | **21.08** | **13.49** | **14.65** | **7.49** |

AIC: Akaike Information Criterion; APP: average posterior probability; BC: black carbon; BIC: Bayesian Information Criterion; $NO_2$: nitrogen dioxide; $PM_{2.5}$: fine particulate matter

**Supplementary Table 4**. Mean levels of fine particulate matter (PM2.5) (µg/m3) in each age by latent classes.

| Age | Low, decreasing (6.6%) | Average-low, decreasing (18.0%) | Average-high, decreasing (53.5%) | High, decreasing (21.9%) |
|---|---|---|---|---|
| | Mean (SD) µg/m$^3$ | | | |
| 0 | 27.41 (2.99) | 28.04 (2.21) | 29.48 (1.78) | 32.50 (1.79) |
| 1 | 26.51 (2.90) | 27.72 (1.71) | 29.13 (1.33) | 29.98 (1.41) |
| 2 | 22.47 (2.84) | 24.07 (2.38) | 25.67 (2.19) | 29.19 (2.22) |
| 3 | 20.97 (2.09) | 22.90 (1.31) | 24.25 (0.92) | 25.36 (0.95) |
| 4 | 22.63 (2.35) | 25.26 (2.09) | 26.73 (2.08) | 26.47 (2.32) |
| 5 | 21.55 (1.68) | 24.47 (1.44) | 26.32 (1.05) | 28.47 (0.94) |
| 6 | 17.31 (2.02) | 19.70 (2.10) | 21.55 (2.44) | 25.27 (2.56) |
| 7 | 15.82 (0.89) | 17.96 (0.98) | 19.47 (0.65) | 20.70 (0.69) |
| 8 | 14.94 (0.94) | 16.93 (0.95) | 18.42 (0.70) | 19.85 (0.71) |
| 9 | 15.88 (1.18) | 18.03 (1.54) | 19.49 (1.37) | 19.58 (1.48) |
| 10 | 15.92 (1.07) | 17.94 (1.16) | 19.57 (0.76) | 20.85 (0.67) |
| 11 | 17.92 (1.72) | 20.27 (2.23) | 21.96 (2.13) | 21.52 (2.25) |
| 12 | 15.70 (1.55) | 17.69 (1.57) | 19.60 (1.88) | 22.73 (1.99) |
| 13 | 14.77 (0.80) | 16.59 (1.07) | 18.15 (0.75) | 19.22 (0.72) |
| 14 | 15.27 (1.00) | 17.12 (1.32) | 18.70 (1.04) | 19.31 (1.06) |
| 15 | 15.03 (0.89) | 16.86 (1.09) | 18.51 (0.80) | 19.76 (0.74) |
| 16 | 12.98 (1.20) | 14.50 (1.21) | 16.05 (1.33) | 18.34 (1.37) |
| 17 | 11.58 (0.82) | 12.90 (0.83) | 14.18 (0.73) | 15.55 (0.75) |
| 18 | 11.83 (0.93) | 13.15 (1.04) | 14.32 (0.87) | 14.75 (0.89) |
| Overall | 17.71 (0.68) | 19.60 (0.50) | 21.13 (0.49) | 22.60 (0.54) |

SD: standard deviation

**Supplementary Table 5**. Mean levels of nitrogen dioxide ($NO_2$) ($\mu g/m^3$) in each age by latent classes.

| Age | Low, decreasing (9.7%) | Average-low, decreasing (17.2%) | Average, decreasing (26.9%) | Average-high, decreasing (30.0%) | High, decreasing (9.4%) | High, decreasing fast (6.8%) |
|---|---|---|---|---|---|---|
| | Mean (SD) $\mu g/m^3$ | | | | | |
| 0 | 44.10 (8.75) | 50.35 (5.27) | 57.40 (4.38) | 63.78 (4.21) | 70.00 (4.60) | 63.88 (6.54) |
| 1 | 40.87 (7.54) | 47.94 (4.51) | 55.01 (3.84) | 61.21 (3.89) | 67.24 (4.15) | 61.16 (6.07) |
| 2 | 35.05 (5.90) | 42.22 (4.13) | 48.73 (4.22) | 54.88 (4.76) | 61.70 (5.16) | 53.59 (7.69) |
| 3 | 33.00 (4.82) | 40.88 (3.84) | 46.83 (3.29) | 51.90 (3.32) | 57.08 (3.79) | 48.91 (7.58) |
| 4 | 35.26 (5.15) | 44.56 (4.73) | 50.91 (4.74) | 55.97 (5.00) | 61.11 (5.87) | 50.32 (10.10) |
| 5 | 34.66 (4.19) | 43.97 (3.71) | 50.70 (3.09) | 56.49 (3.10) | 63.02 (3.44) | 47.25 (10.51) |
| 6 | 28.90 (4.86) | 36.46 (4.49) | 42.38 (4.85) | 48.02 (5.69) | 54.77 (6.27) | 37.88 (9.46) |
| 7 | 26.43 (3.27) | 33.64 (2.71) | 38.57 (2.39) | 42.83 (2.36) | 47.28 (3.08) | 32.20 (7.47) |
| 8 | 24.32 (3.17) | 31.11 (2.81) | 35.58 (2.54) | 39.76 (2.84) | 44.71 (3.11) | 27.71 (5.86) |
| 9 | 26.41 (3.82) | 34.35 (4.33) | 38.73 (4.68) | 42.15 (5.06) | 45.86 (5.80) | 28.70 (6.48) |
| 10 | 25.20 (3.36) | 32.51 (3.17) | 36.87 (2.87) | 41.32 (3.43) | 47.06 (3.40) | 27.41 (5.08) |
| 11 | 25.34 (3.26) | 33.07 (3.42) | 36.91 (3.27) | 40.60 (3.44) | 44.90 (3.86) | 26.36 (4.37) |
| 12 | 23.38 (3.27) | 30.31 (3.18) | 34.03 (2.91) | 38.19 (3.37) | 43.24 (3.58) | 24.35 (3.67) |
| 13 | 22.02 (2.70) | 28.80 (2.87) | 32.01 (2.36) | 35.47 (2.57) | 39.44 (3.08) | 22.67 (3.22) |
| 14 | 21.67 (2.72) | 28.19 (2.83) | 31.38 (2.46) | 34.77 (2.69) | 38.76 (2.93) | 22.27 (3.16) |
| 15 | 21.55 (2.63) | 27.95 (2.87) | 31.08 (2.63) | 34.45 (2.74) | 38.26 (3.15) | 22.13 (3.17) |
| 16 | 20.82 (2.66) | 26.78 (2.85) | 29.93 (2.66) | 33.32 (2.72) | 37.24 (3.03) | 21.45 (3.11) |
| 17 | 18.60 (2.63) | 23.86 (2.86) | 26.69 (2.81) | 29.94 (2.92) | 33.88 (3.29) | 19.34 (3.10) |
| 18 | 19.07 (2.79) | 24.57 (3.18) | 27.28 (3.01) | 29.93 (3.28) | 32.81 (3.72) | 19.52 (3.45) |
| Overall | 27.76 (2.45) | 34.81 (1.68) | 39.54 (1.40) | 43.94 (1.41) | 48.86 (1.96) | 34.63 (3.24) |

SD: standard deviation

**Supplementary Table 6**. Mean levels of black carbon ($10^{-5}$/m) in each age by latent classes.

| Age | Low, decreasing (14.7%) | Average-low, decreasing (22.2%) | Average, decreasing (21.0%) | Average-high, decreasing (21.1%) | High, decreasing (13.5%) | High, decreasing fast (7.5%) |
|---|---|---|---|---|---|---|
| | Mean (SD) $10^{-5}$/m | | | | | |
| 0 | 1.34 (0.33) | 1.47 (0.21) | 1.75 (0.23) | 2.02 (0.22) | 2.22 (0.19) | 2.07 (0.23) |
| 1 | 1.32 (0.30) | 1.48 (0.19) | 1.76 (0.21) | 2.04 (0.21) | 2.24 (0.18) | 2.07 (0.21) |
| 2 | 1.15 (0.24) | 1.33 (0.18) | 1.61 (0.22) | 1.87 (0.23) | 2.06 (0.24) | 1.92 (0.26) |
| 3 | 1.06 (0.18) | 1.26 (0.13) | 1.52 (0.15) | 1.78 (0.14) | 1.96 (0.13) | 1.73 (0.25) |
| 4 | 1.21 (0.21) | 1.47 (0.20) | 1.77 (0.24) | 2.07 (0.25) | 2.28 (0.28) | 1.90 (0.41) |
| 5 | 1.25 (0.16) | 1.54 (0.13) | 1.87 (0.16) | 2.19 (0.16) | 2.41 (0.14) | 1.93 (0.43) |
| 6 | 1.04 (0.18) | 1.30 (0.20) | 1.59 (0.24) | 1.85 (0.27) | 2.05 (0.28) | 1.58 (0.41) |
| 7 | 0.92 (0.10) | 1.15 (0.10) | 1.40 (0.11) | 1.62 (0.12) | 1.80 (0.10) | 1.29 (0.33) |
| 8 | 0.97 (0.11) | 1.22 (0.12) | 1.48 (0.14) | 1.72 (0.16) | 1.90 (0.16) | 1.25 (0.33) |
| 9 | 1.16 (0.16) | 1.47 (0.19) | 1.77 (0.23) | 2.05 (0.26) | 2.27 (0.27) | 1.40 (0.37) |
| 10 | 1.15 (0.12) | 1.45 (0.14) | 1.77 (0.17) | 2.04 (0.18) | 2.29 (0.17) | 1.36 (0.28) |
| 11 | 1.24 (0.16) | 1.57 (0.19) | 1.88 (0.21) | 2.18 (0.24) | 2.42 (0.25) | 1.38 (0.25) |
| 12 | 1.10 (0.18) | 1.39 (0.22) | 1.69 (0.26) | 1.95 (0.28) | 2.19 (0.31) | 1.26 (0.25) |
| 13 | 1.02 (0.11) | 1.29 (0.12) | 1.55 (0.12) | 1.78 (0.15) | 2.00 (0.12) | 1.13 (0.17) |
| 14 | 1.06 (0.12) | 1.34 (0.14) | 1.61 (0.17) | 1.85 (0.21) | 2.08 (0.20) | 1.16 (0.16) |
| 15 | 1.00 (0.11) | 1.26 (0.14) | 1.52 (0.15) | 1.74 (0.17) | 1.96 (0.15) | 1.11 (0.16) |
| 16 | 0.92 (0.09) | 1.15 (0.12) | 1.39 (0.13) | 1.59 (0.16) | 1.80 (0.13) | 1.02 (0.14) |
| 17 | 0.85 (0.09) | 1.07 (0.11) | 1.29 (0.13) | 1.47 (0.16) | 1.67 (0.13) | 0.95 (0.14) |
| 18 | 0.94 (0.13) | 1.19 (0.17) | 1.42 (0.19) | 1.62 (0.22) | 1.82 (0.21) | 1.03 (0.18) |
| Overall | 1.09 (0.08) | 1.34 (0.07) | 1.61 (0.07) | 1.86 (0.07) | 2.07 (0.08) | 1.45 (0.13) |

SD: standard deviation

**Supplementary Table 7**. Fine particulate matter ($PM_{2.5}$) classes distribution by socioeconomic indicators.

| **Indicator** | **Low, decreasing (6.1%)** | **Average-low, decreasing (18.0%)** | **Average-high, decreasing (53.5%)** | **High, decreasing (21.9%)** |
|---|---|---|---|---|
| | | | **%** | |
| Maternal education | p<0.001 | | | |
| Low | 2.6 | 12.9 | 58.8 | 25.7 |
| Medium | 6.0 | 17.5 | 53.5 | 23.0 |
| High | 11.3 | 22.5 | 50.6 | 15.6 |
| Area deprivation | p<0.001 | | | |
| 1st Q (least deprived) | 6.6 | 25.7 | 48.3 | 19.4 |
| 2nd quintile | 11.2 | 22.1 | 51.4 | 15.3 |
| 3rd quintile | 8.0 | 17.3 | 52.3 | 22.4 |
| 4th quintile | 3.6 | 11.7 | 60.9 | 23.8 |
| 5th Q (most deprived) | 2.9 | 11.7 | 56.0 | 29.4 |
| Moving status from pregnancy to 18 years | p<0.001 | | | |
| Moved address | 26.4 | 37.1 | 30.7 | 5.8 |

$PM_{2.5}$: fine particulate matter; Q: quintile

**Supplementary Table 8**. Nitrogen dioxide ($NO_2$) classes distribution by socioeconomic indicators.

| | Air pollutant classes | | | | | |
|---|---|---|---|---|---|---|
| | **Low, decreasing (9.7%)** | **Average-low, decreasing (17.2%)** | **Average, decreasing (26.9%)** | **Average-high, decreasing (30.0%)** | **High, decreasing (9.4%)** | **High, decreasing fast (6.8%)** |
| **Indicator** | % | | | | | |
| Maternal education | p<0.001 | | | | | |
| Low | 4.7 | 17.1 | 33.5 | 33.7 | 8.4 | 2.6 |
| Medium | 9.8 | 18.1 | 28.3 | 28.5 | 8.8 | 6.5 |
| High | 12.4 | 14.2 | 17.8 | 33.2 | 12.3 | 10.1 |
| Area deprivation | p<0.001 | | | | | |
| 1st Q (least deprived) | 11.4 | 25.3 | 28.0 | 26.6 | 3.3 | 5.4 |
| 2nd quintile | 20.2 | 21.9 | 22.8 | 23.0 | 5.2 | 6.9 |
| 3rd quintile | 10.5 | 14.2 | 25.9 | 30.4 | 10.7 | 8.3 |
| 4th quintile | 3.4 | 11.3 | 24.9 | 39.6 | 14.9 | 5.9 |
| 5th Q (most deprived) | 1.6 | 12.7 | 33.3 | 31.1 | 14.2 | 7.1 |
| Moving status from pregnancy to 18 years | p<0.001 | | | | | |
| Moved address | 21.1 | 18.8 | 17.8 | 10.7 | 4.3 | 27.2 |

Q: quintile

**Supplementary Table 9**. Black carbon (BC) classes distribution by socioeconomic indicators.

| | Air pollutant classes | | | | | |
|---|---|---|---|---|---|---|
| | Low, decreasing (14.7%) | Average-low, decreasing (22.2%) | Average, decreasing (21.0%) | Average-high, decreasing (21.1%) | High, decreasing (13.5%) | High, decreasing fast (7.5%) |
| **Indicator** | % | | | | | |
| Maternal education | p<0.001 | | | | | |
| Low | 7.0 | 27.4 | 26.2 | 21.6 | 14.0 | 3.8 |
| Medium | 14.7 | 23.1 | 22.1 | 20.3 | 12.9 | 6.9 |
| High | 19.2 | 15.9 | 14.2 | 23.5 | 15.4 | 11.8 |
| Area deprivation | p<0.001 | | | | | |
| 1st Q (least deprived) | 22.2 | 29.5 | 26.1 | 14.7 | 2.4 | 5.1 |
| 2nd quintile | 25.0 | 27.1 | 16.5 | 18.0 | 6.5 | 6.9 |
| 3rd quintile | 14.6 | 21.3 | 16.2 | 24.8 | 14.0 | 9.1 |
| 4th quintile | 6.1 | 14.5 | 22.6 | 24.9 | 24.3 | 7.6 |
| 5th Q (most deprived) | 3.5 | 17.9 | 24.8 | 23.0 | 22.1 | 8.7 |
| Moving status from pregnancy to 18 years | p<0.001 | | | | | |
| Moved address | 34.3 | 20.8 | 9.7 | 8.6 | 2.6 | 23.9 |

Q: quintile

**Supplementary Table 10.** Association between fine particulate matter ($PM_{2.5}$) from pregnancy to 18 years with cardiovascular health indicators at age 18 years.

| Outcome | Exposure | β (95% CI) | p-value |
|---|---|---|---|
| Peripheral SBP, mmHg | $PM_{2.5}$ from pregnancy-18y | 0.14 (-0.30, 0.58) | 0.541 |
| | Low, decreasing | Reference | 0.991 |
| | Average-low, decreasing | 0.25 (-1.39, 1.89) | |
| | Average-high, decreasing | 0.13 (-1.37, 1.62) | |
| | High, decreasing | 0.18 (-1.44, 1.79) | |
| Central SBP, mmHg | $PM_{2.5}$ from pregnancy-18y | 0.31 (-0.07, 0.68) | 0.108 |
| | Low, decreasing | Reference | 0.675 |
| | Average-low, decreasing | 0.52 (-0.87, 1.91) | |
| | Average-high, decreasing | 0.53 (-0.73, 1.80) | |
| | High, decreasing | 0.84 (-0.53, 2.21) | |
| Peripheral DBP, mmHg | $PM_{2.5}$ from pregnancy-18y | 0.46 (0.14, 0.78) | 0.005 |
| | Low, decreasing | Reference | 0.238 |
| | Average-low, decreasing | 0.61 (-0.57, 1.80) | |
| | Average-high, decreasing | 0.69 (-0.39, 1.77) | |
| | High, decreasing | 1.16 (-0.01, 2.33) | |
| Central DBP, mmHg | $PM_{2.5}$ from pregnancy-18y | 0.50 (0.17, 0.83) | 0.003 |
| | Low, decreasing | Reference | 0.195 |
| | Average-low, decreasing | 0.68 (-0.54, 1.89) | |
| | Average-high, decreasing | 0.81 (-0.30, 1.91) | |
| | High, decreasing | 1.26 (0.07, 2.46) | |
| CIMT, µm | $PM_{2.5}$ from pregnancy-18y | -2.28 (-4.03, -0.52) | 0.011 |
| | Low, decreasing | Reference | 0.103 |
| | Average-low, decreasing | -3.29 (-9.70, 3.11) | |
| | Average-high, decreasing | -4.90 (-10.69, 0.90) | |
| | High, decreasing | -7.39 (-13.68, -1.10) | |
| PWV, m/s | $PM_{2.5}$ from pregnancy-18y | 0.02 (-0.01, 0.05) | 0.117 |
| | Low, decreasing | Reference | 0.400 |
| | Average-low, decreasing | 0.04 (-0.06, 0.14) | |
| | Average-high, decreasing | 0.07 (-0.02, 0.17) | |
| | High, decreasing | 0.06 (-0.04, 0.16) | |
| Heart rate, bpm | $PM_{2.5}$ from pregnancy-18y | 0.31 (-0.14, 0.76) | 0.180 |
| | Low, decreasing | Reference | 0.468 |
| | Average-low, decreasing | 0.26 (-1.41, 1.94) | |
| | Average-high, decreasing | 0.89 (-0.64, 2.41) | |
| | High, decreasing | 0.44 (-1.21, 2.09) | |
| AIx, % | $PM_{2.5}$ from pregnancy-18y | -0.09 (-0.65, 0.48) | 0.765 |
| | Low, decreasing | Reference | 0.699 |
| | Average-low, decreasing | 0.07 (-2.02, 2.16) | |
| | Average-high, decreasing | -0.23 (-2.14, 1.67) | |
| | High, decreasing | 0.49 (-1.57, 2.55) | |

AIx: augmentation index; CIMT: carotid media-intima thickness; DBP: diastolic blood pressure; $PM_{2.5}$: fine particulate matter; PWV: pulse wave velocity; SBP: systolic blood pressure

**Supplementary Table 11.** Association between nitrogen dioxide ($NO_2$) from pregnancy to 18 years with cardiovascular health indicators at age 18 years.

| Outcome | Exposure | β (95% CI) | p-value |
|---|---|---|---|
| Peripheral SBP, mmHg | $NO_2$ from pregnancy-18y | 0.15 (-0.35, 0.64) | 0.565 |
| | Low, decreasing | Reference | 0.315 |
| | Average-low, decreasing | -0.42 (-1.87, 1.03) | |
| | Average, decreasing | 0.36 (-1.00, 1.73) | |
| | Average-high, decreasing | -0.33 (-1.67, 1.01) | |
| | High, decreasing | 0.57 (-1.11, 2.25) | |
| | High, decreasing fast | 1.10 (-0.71, 2.90) | |
| Central SBP, mmHg | $NO_2$ from pregnancy-18y | 0.30 (-0.12, 0.72) | 0.161 |
| | Low, decreasing | Reference | 0.249 |
| | Average-low, decreasing | 0.17 (-1.06, 1.41) | |
| | Average, decreasing | 0.67 (-0.49, 1.83) | |
| | Average-high, decreasing | 0.24 (-0.90, 1.38) | |
| | High, decreasing | 1.21 (-0.22, 2.64) | |
| | High, decreasing fast | 1.37 (-0.17, 2.90) | |
| Peripheral DBP, mmHg | $NO_2$ from pregnancy-18y | 0.45 (0.09, 0.81) | 0.015 |
| | Low, decreasing | Reference | 0.108 |
| | Average-low, decreasing | 0.85 (-0.21, 1.90) | |
| | Average, decreasing | 0.86 (-0.14, 1.85) | |
| | Average-high, decreasing | 0.83 (-0.15, 1.81) | |
| | High, decreasing | 1.73 (0.50, 2.95) | |
| | High, decreasing fast | 1.47 (0.16, 2.79) | |
| Central DBP, mmHg | $NO_2$ from pregnancy-18y | 0.50 (0.13, 0.87) | 0.009 |
| | Low, decreasing | Reference | 0.095 |
| | Average-low, decreasing | 0.86 (-0.22, 1.94) | |
| | Average, decreasing | 0.94 (-0.08, 1.96) | |
| | Average-high, decreasing | 1.00 (0.00, 1.99) | |
| | High, decreasing | 1.78 (0.52, 3.03) | |
| | High, decreasing fast | 1.61 (0.26, 2.95) | |
| CIMT, µm | $NO_2$ from pregnancy-18y | -0.42 (-2.41, 1.57) | 0.681 |
| | Low, decreasing | Reference | 0.835 |
| | Average-low, decreasing | -1.64 (-7.41, 4.14) | |
| | Average, decreasing | -1.38 (-6.85, 4.10) | |
| | Average-high, decreasing | -0.46 (-5.86, 4.93) | |
| | High, decreasing | -3.95 (-10.71, 2.80) | |
| | High, decreasing fast | 0.07 (-7.16, 7.29) | |
| PWV, m/s | $NO_2$ from pregnancy-18y | 0.01 (-0.02, 0.04) | 0.659 |
| | Low, decreasing | Reference | 0.323 |
| | Average-low, decreasing | 0.08 (-0.01, 0.17) | |
| | Average, decreasing | 0.08 (0.00, 0.17) | |
| | Average-high, decreasing | 0.05 (-0.03, 0.14) | |
| | High, decreasing | 0.02 (-0.09, 0.13) | |
| | High, decreasing fast | 0.02 (-0.09, 0.14) | |
| Heart rate, bpm | $NO_2$ from pregnancy-18y | 0.18 (-0.33, 0.68) | 0.500 |
| | Low, decreasing | Reference | 0.195 |

| | | | |
|---|---|---|---|
| | Average-low, decreasing | 0.65 (-0.83, 2.14) | |
| | Average, decreasing | 0.49 (-0.91, 1.89) | |
| | Average-high, decreasing | 1.18 (-0.20, 2.55) | |
| | High, decreasing | -0.57 (-2.29, 1.16) | |
| | High, decreasing fast | 0.41 (-1.44, 2.26) | |
| AIx, % | $NO_2$ from pregnancy-18y | 0.02 (-0.62, 0.65) | 0.956 |
| | Low, decreasing | Reference | 0.823 |
| | Average-low, decreasing | 0.60 (-1.25, 2.45) | |
| | Average, decreasing | 0.68 (-1.07, 2.43) | |
| | Average-high, decreasing | -0.04 (-1.76, 1.67) | |
| | High, decreasing | 0.84 (-1.31, 2.98) | |
| | High, decreasing fast | 0.09 (-2.21, 2.39) | |

AIx: augmentation index; CIMT: carotid media-intima thickness; DBP: diastolic blood pressure; $NO_2$: nitrogen dioxide; PWV: pulse wave velocity; SBP: systolic blood pressure

**Supplementary Table 12.** Association between black carbon (BC) from pregnancy to 18 years with cardiovascular health indicators at age 18 years.

| Outcome | Exposure | β (95% CI) | p-value |
|---|---|---|---|
| Peripheral SBP, mmHg | BC from pregnancy-18y | 0.10 (-0.56, 0.76) | 0.772 |
| | Low, decreasing | Reference | 0.063 |
| | Average-low, decreasing | 0.26 (-0.98, 1.50) | |
| | Average, decreasing | 0.30 (-0.95, 1.55) | |
| | Average-high, decreasing | 1.13 (-0.14, 2.40) | |
| | High, decreasing | 0.14 (-1.30, 1.58) | |
| | High, decreasing fast | 2.18 (0.54, 3.83) | |
| Central SBP, mmHg | BC from pregnancy-18y | 0.18 (-0.38, 0.74) | 0.529 |
| | Low, decreasing | Reference | 0.138 |
| | Average-low, decreasing | 0.52 (-0.53, 1.58) | |
| | Average, decreasing | 0.57 (-0.49, 1.64) | |
| | Average-high, decreasing | 1.10 (0.02, 2.18) | |
| | High, decreasing | 0.28 (-0.94, 1.51) | |
| | High, decreasing fast | 1.72 (0.32, 3.12) | |
| Peripheral DBP, mmHg | BC from pregnancy-18y | 0.32 (-0.16, 0.80) | 0.195 |
| | Low, decreasing | Reference | 0.154 |
| | Average-low, decreasing | 0.89 (-0.01, 1.80) | |
| | Average, decreasing | 0.79 (-0.12, 1.70) | |
| | Average-high, decreasing | 1.00 (0.08, 1.93) | |
| | High, decreasing | 0.89 (-0.16, 1.94) | |
| | High, decreasing fast | 1.59 (0.39, 2.79) | |
| Central DBP, mmHg | BC from pregnancy-18y | 0.41 (-0.09, 0.90) | 0.106 |
| | Low, decreasing | Reference | 0.102 |
| | Average-low, decreasing | 0.95 (0.03, 1.88) | |
| | Average, decreasing | 0.90 (-0.03, 1.83) | |
| | Average-high, decreasing | 1.17 (0.22, 2.12) | |
| | High, decreasing | 1.03 (-0.04, 2.11) | |
| | High, decreasing fast | 1.70 (0.47, 2.93) | |
| CIMT, µm | BC from pregnancy-18y | 1.06 (-1.58, 3.70) | 0.431 |
| | Low, decreasing | Reference | 0.189 |
| | Average-low, decreasing | -3.23 (-8.12, 1.65) | |
| | Average, decreasing | -2.33 (-7.31, 2.65) | |
| | Average-high, decreasing | 1.99 (-3.04, 7.03) | |
| | High, decreasing | -0.86 (-6.64, 4.93) | |
| | High, decreasing fast | -4.09 (-10.66, 2.48) | |
| PWV, m/s | BC from pregnancy-18y | -0.02 (-0.06, 0.02) | 0.336 |
| | Low, decreasing | Reference | 0.005 |
| | Average-low, decreasing | 0.14 (0.06, 0.21) | |
| | Average, decreasing | 0.06 (-0.02, 0.14) | |
| | Average-high, decreasing | 0.03 (-0.05, 0.11) | |
| | High, decreasing | 0.03 (-0.06, 0.12) | |
| | High, decreasing fast | 0.00 (-0.10, 0.11) | |
| Heart rate, bpm | BC from pregnancy-18y | 0.68 (0.00, 1.36) | 0.049 |
| | Low, decreasing | Reference | 0.159 |

| | | | |
|---|---|---|---|
| | Average-low, decreasing | 0.84 (-0.43, 2.11) | |
| | Average, decreasing | 1.19 (-0.09, 2.47) | |
| | Average-high, decreasing | 1.19 (-0.12, 2.49) | |
| | High, decreasing | 1.76 (0.28, 3.24) | |
| | High, decreasing fast | 1.99 (0.30, 3.68) | |
| AIx, % | BC from pregnancy-18y | -0.30 (-1.15, 0.54) | 0.479 |
| | Low, decreasing | Reference | 0.546 |
| | Average-low, decreasing | -0.60 (-2.18, 0.97) | |
| | Average, decreasing | -0.32 (-1.91, 1.27) | |
| | Average-high, decreasing | -0.25 (-1.87, 1.37) | |
| | High, decreasing | -1.28 (-3.11, 0.55) | |
| | High, decreasing fast | -1.64 (-3.73, 0.46) | |

AIx: augmentation index; BC: black carbon; CIMT: carotid media-intima thickness; DBP: diastolic blood pressure; PWV: pulse wave velocity; SBP: systolic blood pressure

**Supplementary Table 13.** Association between air pollution at different age ranges and cardiovascular health markers at age 18 years.

| Outcome | Exposure | 0-5 years beta (95%CI) | 0-5 years p-value | 6-12 years beta (95%CI) | 6-12 years p-value | 13-18 years beta (95%CI) | 13-18 years p-value | p-value difference by age |
|---|---|---|---|---|---|---|---|---|
| Peripheral SBP, mmHg | $PM_{2.5}$ (µg/m$^3$) | 0.10 (-0.09, 0.28) | 0.303 | 0.09 (-0.16, 0.34) | 0.487 | 0.01 (-0.27, 0.29) | 0.962 | 0.488 |
| | $NO_2$ (µg/m$^3$) | 0.01 (-0.03, 0.06) | 0.546 | 0.02 (-0.04, 0.08) | 0.585 | 0.00 (-0.07, 0.07) | 0.996 | 0.644 |
| | BC ($10^{-5}$/m) | 0.17 (-0.94, 1.29) | 0.757 | 0.09 (-0.99, 1.17) | 0.873 | 0.09 (-1.07, 1.26) | 0.876 | 0.977 |
| Central SBP, mmHg | $PM_{2.5}$ (µg/m$^3$) | 0.19 (0.04, 0.35) | 0.015 | 0.20 (-0.01, 0.41) | 0.066 | 0.10 (-0.14, 0.34) | 0.418 | 0.190 |
| | $NO_2$ (µg/m$^3$) | 0.03 (-0.01, 0.07) | 0.104 | 0.04 (-0.01, 0.09) | 0.127 | 0.02 (-0.04, 0.08) | 0.514 | 0.372 |
| | BC ($10^{-5}$/m) | 0.47 (-0.47, 1.42) | 0.328 | 0.28 (-0.63, 1.20) | 0.545 | 0.28 (-0.72, 1.27) | 0.585 | 0.879 |
| Peripheral DBP, mmHg | $PM_{2.5}$ (µg/m$^3$) | 0.27 (0.13, 0.40) | <0.001 | 0.26 (0.08, 0.44) | 0.005 | 0.20 (0.00, 0.41) | 0.051 | 0.432 |
| | $NO_2$ (µg/m$^3$) | 0.05 (0.02, 0.08) | 0.003 | 0.05 (0.01, 0.10) | 0.014 | 0.04 (-0.01, 0.10) | 0.084 | 0.712 |
| | BC ($10^{-5}$/m) | 1.01 (0.21, 1.82) | 0.014 | 0.43 (-0.35, 1.21) | 0.283 | 0.54 (-0.31, 1.39) | 0.210 | 0.146 |
| Central DBP, mmHg | $PM_{2.5}$ (µg/m$^3$) | 0.28 (0.14, 0.42) | <0.001 | 0.28 (0.10, 0.47) | 0.003 | 0.23 (0.02, 0.43) | 0.033 | 0.455 |
| | $NO_2$ (µg/m$^3$) | 0.05 (0.02, 0.09) | 0.002 | 0.06 (0.02, 0.10) | 0.008 | 0.05 (0.00, 0.10) | 0.065 | 0.669 |
| | BC ($10^{-5}$/m) | 1.18 (0.35, 2.01) | 0.005 | 0.56 (-0.25, 1.36) | 0.175 | 0.67 (-0.20, 1.54) | 0.130 | 0.124 |
| CIMT, µm | $PM_{2.5}$ (µg/m$^3$) | -1.36 (-2.09, -0.63) | <0.001 | -1.16 (-2.15, -0.18) | 0.021 | -1.02 (-2.12, 0.09) | 0.072 | 0.778 |
| | $NO_2$ (µg/m$^3$) | -0.09 (-0.27, 0.09) | 0.336 | -0.08 (-0.31, 0.16) | 0.525 | -0.02 (-0.30, 0.26) | 0.907 | 0.644 |
| | BC ($10^{-5}$/m) | -0.39 (-4.8, 4.01) | 0.861 | 1.48 (-2.83, 5.79) | 0.501 | 1.97 (-2.7, 6.64) | 0.409 | 0.498 |
| PWV, m/s | $PM_{2.5}$ (µg/m$^3$) | 0.00 (-0.01, 0.01) | 0.913 | 0.01 (0.00, 0.03) | 0.069 | 0.02 (0.00, 0.03) | 0.097 | 0.090 |
| | $NO_2$ (µg/m$^3$) | 0.00 (0.00, 0.00) | 0.419 | 0.00 (0.00, 0.01) | 0.474 | 0.00 (0.00, 0.01) | 0.210 | 0.102 |
| | BC ($10^{-5}$/m) | -0.06 (-0.13, 0.01) | 0.105 | -0.03 (-0.10, 0.04) | 0.427 | -0.01 (-0.08, 0.07) | 0.843 | 0.260 |
| Heart rate, bpm | $PM_{2.5}$ (µg/m$^3$) | 0.02 (-0.17, 0.20) | 0.874 | 0.10 (-0.15, 0.35) | 0.443 | 0.20 (-0.09, 0.48) | 0.180 | 0.257 |
| | $NO_2$ (µg/m$^3$) | 0.01 (-0.03, 0.06) | 0.589 | 0.01 (-0.05, 0.07) | 0.792 | 0.02 (-0.05, 0.10) | 0.503 | 0.583 |
| | BC ($10^{-5}$/m) | 1.14 (0.00, 2.28) | 0.049 | 0.84 (-0.26, 1.95) | 0.135 | 0.97 (-0.22, 2.16) | 0.109 | 0.661 |
| AIx, % | $PM_{2.5}$ (µg/m$^3$) | 0.06 (-0.17, 0.30) | 0.601 | 0.02 (-0.30, 0.34) | 0.902 | -0.11 (-0.46, 0.25) | 0.562 | 0.340 |
| | $NO_2$ (µg/m$^3$) | 0.00 (-0.06, 0.06) | 0.986 | 0.02 (-0.06, 0.09) | 0.635 | 0.00 (-0.09, 0.09) | 0.996 | 0.535 |
| | BC ($10^{-5}$/m) | -0.44 (-1.86, 0.97) | 0.538 | -0.21 (-1.58, 1.17) | 0.769 | -0.45 (-1.93, 1.04) | 0.553 | 0.647 |

AIx: augmentation index; BC: black carbon; CIMT: carotid media-intima thickness; DBP: diastolic blood pressure; $NO_2$: nitrogen dioxide; $PM_{2.5}$: fine particulate matter; PWV: pulse wave velocity; SBP: systolic blood pressure

**Supplementary Table 14**. Association between fine particulate matter ($PM_{2.5}$) and cardiovascular health markers at 18 years in males and females.

| Outcome | Exposure | Males beta (95%CI) | p-value | Females beta (95%CI) | p-value | p-value sex interaction |
|---|---|---|---|---|---|---|
| Peripheral SBP, mmHg | $PM_{2.5}$ from pregnancy-18y | 0.14 (-0.57, 0.86) | 0.693 | 0.13 (-0.43, 0.68) | 0.657 | 0.671 |
| | Low, decreasing | Reference | 0.638 | Reference | 0.867 | 0.871 |
| | Average-low, decreasing | 1.58 (-1.07, 4.23) | | -0.77 (-2.84, 1.29) | | |
| | Average-high, decreasing | 0.77 (-1.66, 3.20) | | -0.34 (-2.20, 1.53) | | |
| | High, decreasing | 0.77 (-1.84, 3.38) | | -0.26 (-2.29, 1.77) | | |
| Central SBP, mmHg | $PM_{2.5}$ from pregnancy-18y | 0.28 (-0.30, 0.86) | 0.342 | 0.32 (-0.17, 0.82) | 0.204 | 0.799 |
| | Low, decreasing | Reference | 0.449 | Reference | 0.568 | 0.824 |
| | Average-low, decreasing | 1.47 (-0.67, 3.62) | | -0.24 (-2.08, 1.59) | | |
| | Average-high, decreasing | 0.66 (-1.31, 2.62) | | 0.46 (-1.20, 2.12) | | |
| | High, decreasing | 1.10 (-1.02, 3.22) | | 0.64 (-1.17, 2.44) | | |
| Peripheral DBP, mmHg | $PM_{2.5}$ from pregnancy-18y | 0.46 (-0.04, 0.95) | 0.071 | 0.45 (0.03, 0.87) | 0.037 | 0.837 |
| | Low, decreasing | Reference | 0.556 | Reference | 0.272 | 0.522 |
| | Average-low, decreasing | 0.97 (-0.87, 2.81) | | 0.31 (-1.24, 1.87) | | |
| | Average-high, decreasing | 0.49 (-1.20, 2.18) | | 0.84 (-0.57, 2.24) | | |
| | High, decreasing | 1.01 (-0.81, 2.82) | | 1.25 (-0.28, 2.78) | | |
| Central DBP, mmHg | $PM_{2.5}$ from pregnancy-18y | 0.52 (0.01, 1.03) | 0.044 | 0.47 (0.04, 0.90) | 0.031 | 0.757 |
| | Low, decreasing | Reference | 0.584 | Reference | 0.262 | 0.601 |
| | Average-low, decreasing | 1.01 (-0.88, 2.90) | | 0.40 (-1.19, 1.99) | | |
| | Average-high, decreasing | 0.66 (-1.07, 2.39) | | 0.92 (-0.52, 2.35) | | |
| | High, decreasing | 1.15 (-0.71, 3.01) | | 1.33 (-0.23, 2.90) | | |
| CIMT, µm | $PM_{2.5}$ from pregnancy-18y | -1.55 (-4.31, 1.20) | 0.269 | -2.80 (-5.07, -0.53) | 0.016 | 0.347 |
| | Low, decreasing | Reference | 0.718 | Reference | 0.026 | 0.138 |
| | Average-low, decreasing | 3.57 (-6.5, 13.65) | | -8.30 (-16.58, -0.03) | | |
| | Average-high, decreasing | 0.29 (-8.92, 9.50) | | -8.44 (-15.87, -1.02) | | |
| | High, decreasing | -0.28 (-10.25, 9.68) | | -12.46 (-20.54, -4.37) | | |
| PWV, m/s | $PM_{2.5}$ from pregnancy-18y | 0.02 (-0.02, 0.07) | 0.314 | 0.02 (-0.02, 0.05) | 0.282 | 0.666 |
| | Low, decreasing | Reference | 0.661 | Reference | 0.349 | 0.894 |

| | | | | | | |
|---|---|---|---|---|---|---|
| | Average-low, decreasing | 0.10 (-0.07, 0.27) | | -0.01 (-0.14, 0.11) | | |
| | Average-high, decreasing | 0.09 (-0.06, 0.24) | | 0.06 (-0.06, 0.17) | | |
| | High, decreasing | 0.08 (-0.09, 0.24) | | 0.04 (-0.08, 0.17) | | |
| Heart rate, bpm | $PM_{2.5}$ from pregnancy-18y | 0.60 (-0.10, 1.29) | 0.092 | 0.10 (-0.49, 0.70) | 0.740 | 0.409 |
| | Low, decreasing | Reference | 0.055 | Reference | 0.945 | 0.267 |
| | Average-low, decreasing | 0.23 (-2.36, 2.81) | | 0.35 (-1.85, 2.55) | | |
| | Average-high, decreasing | 2.10 (-0.27, 4.47) | | -0.03 (-2.02, 1.95) | | |
| | High, decreasing | 1.19 (-1.36, 3.74) | | -0.11 (-2.27, 2.05) | | |
| AIx, % | $PM_{2.5}$ from pregnancy-18y | -0.07 (-0.88, 0.74) | 0.862 | -0.11 (-0.89, 0.67) | 0.776 | 0.896 |
| | Low, decreasing | Reference | 0.140 | Reference | 0.876 | 0.677 |
| | Average-low, decreasing | 0.41 (-2.57, 3.40) | | -0.29 (-3.2, 2.63) | | |
| | Average-high, decreasing | -1.17 (-3.91, 1.57) | | 0.45 (-2.19, 3.08) | | |
| | High, decreasing | 0.49 (-2.46, 3.44) | | 0.41 (-2.46, 3.28) | | |

AIx: augmentation index; CIMT: carotid media-intima thickness; DBP: diastolic blood pressure; $PM_{2.5}$: fine particulate matter; PWV: pulse wave velocity; SBP: systolic blood pressure

**Supplementary Table 15.** Association between nitrogen dioxide ($NO_2$) and cardiovascular health markers at 18 years in males and females.

| Outcome | Exposure | Males | | Females | | p-value sex |
|---|---|---|---|---|---|---|
| | | beta (95%CI) | p-value | beta (95%CI) | p-value | interaction |
| Peripheral SBP, mmHg | $NO_2$ from pregnancy-18y | 0.64 (-0.14, 1.42) | 0.108 | -0.24 (-0.88, 0.40) | 0.460 | 0.034 |
| | Low, decreasing | Reference | 0.606 | Reference | 0.255 | 0.862 |
| | Average-low, decreasing | -0.35 (-2.61, 1.91) | | -0.42 (-2.30, 1.46) | | |
| | Average, decreasing | 1.00 (-1.13, 3.14) | | -0.15 (-1.92, 1.62) | | |
| | Average-high, decreasing | 0.02 (-2.09, 2.13) | | -0.61 (-2.34, 1.12) | | |
| | High, decreasing | 1.25 (-1.38, 3.89) | | 0.04 (-2.14, 2.22) | | |
| | High, decreasing fast | 0.16 (-2.60, 2.93) | | 1.98 (-0.40, 4.36) | | |
| Central SBP, mmHg | $NO_2$ from pregnancy-18y | 0.60 (-0.03, 1.23) | 0.062 | 0.07 (-0.50, 0.64) | 0.807 | 0.095 |
| | Low, decreasing | Reference | 0.765 | Reference | 0.327 | 0.714 |
| | Average-low, decreasing | 0.10 (-1.73, 1.93) | | 0.28 (-1.40, 1.95) | | |
| | Average, decreasing | 0.93 (-0.80, 2.66) | | 0.46 (-1.11, 2.04) | | |
| | Average-high, decreasing | 0.55 (-1.15, 2.26) | | 0.00 (-1.54, 1.55) | | |
| | High, decreasing | 1.25 (-0.88, 3.38) | | 1.19 (-0.75, 3.13) | | |
| | High, decreasing fast | 0.81 (-1.43, 3.04) | | 1.86 (-0.25, 3.98) | | |
| Peripheral DBP, mmHg | $NO_2$ from pregnancy-18y | 0.69 (0.15, 1.23) | 0.013 | 0.26 (-0.23, 0.74) | 0.300 | 0.125 |
| | Low, decreasing | Reference | 0.440 | Reference | 0.333 | 0.324 |
| | Average-low, decreasing | 0.53 (-1.04, 2.10) | | 1.13 (-0.29, 2.55) | | |
| | Average, decreasing | 1.09 (-0.40, 2.58) | | 0.66 (-0.68, 2.00) | | |
| | Average-high, decreasing | 1.04 (-0.43, 2.50) | | 0.67 (-0.64, 1.98) | | |
| | High, decreasing | 1.78 (-0.05, 3.62) | | 1.65 (0.00, 3.30) | | |
| | High, decreasing fast | 1.41 (-0.52, 3.33) | | 1.49 (-0.31, 3.29) | | |
| Central DBP, mmHg | $NO_2$ from pregnancy-18y | 0.79 (0.23, 1.35) | 0.006 | 0.26 (-0.23, 0.76) | 0.296 | 0.088 |
| | Low, decreasing | Reference | 0.297 | Reference | 0.356 | 0.256 |
| | Average-low, decreasing | 0.56 (-1.05, 2.17) | | 1.13 (-0.33, 2.59) | | |
| | Average, decreasing | 1.29 (-0.23, 2.81) | | 0.65 (-0.72, 2.02) | | |
| | Average-high, decreasing | 1.35 (-0.16, 2.85) | | 0.73 (-0.61, 2.07) | | |
| | High, decreasing | 1.90 (0.02, 3.78) | | 1.65 (-0.04, 3.33) | | |

| | | | | | | |
|---|---|---|---|---|---|---|
| | High, decreasing fast | 1.62 (-0.35, 3.59) | | 1.55 (-0.28, 3.39) | | |
| CIMT, µm | $NO_2$ from pregnancy-18y | 0.89 (-2.16, 3.94) | 0.567 | -1.48 (-4.11, 1.16) | 0.272 | 0.288 |
| | Low, decreasing | Reference | 0.755 | Reference | 0.112 | 0.120 |
| | Average-low, decreasing | 1.52 (-7.20, 10.24) | | -4.37 (-12.08, 3.34) | | |
| | Average, decreasing | 0.90 (-7.44, 9.24) | | -3.27 (-10.53, 3.98) | | |
| | Average-high, decreasing | -1.06 (-9.29, 7.16) | | -0.22 (-7.36, 6.92) | | |
| | High, decreasing | -0.54 (-10.80, 9.71) | | -6.76 (-15.73, 2.22) | | |
| | High, decreasing fast | -6.11 (-17.07, 4.86) | | 5.35 (-4.24, 14.95) | | |
| PWV, m/s | $NO_2$ from pregnancy-18y | 0.01 (-0.04, 0.06) | 0.578 | 0.00 (-0.04, 0.04) | 0.960 | 0.369 |
| | Low, decreasing | Reference | 0.757 | Reference | 0.405 | 0.265 |
| | Average-low, decreasing | 0.05 (-0.09, 0.19) | | 0.10 (-0.02, 0.21) | | |
| | Average, decreasing | 0.10 (-0.03, 0.23) | | 0.07 (-0.04, 0.18) | | |
| | Average-high, decreasing | 0.04 (-0.09, 0.18) | | 0.06 (-0.05, 0.16) | | |
| | High, decreasing | 0.04 (-0.13, 0.20) | | 0.00 (-0.14, 0.13) | | |
| | High, decreasing fast | 0.04 (-0.14, 0.22) | | 0.00 (-0.15, 0.15) | | |
| Heart rate, bpm | $NO_2$ from pregnancy-18y | 0.53 (-0.23, 1.30) | 0.171 | -0.13 (-0.82, 0.55) | 0.702 | 0.232 |
| | Low, decreasing | Reference | 0.246 | Reference | 0.593 | 0.148 |
| | Average-low, decreasing | 0.54 (-1.68, 2.76) | | 0.74 (-1.26, 2.74) | | |
| | Average, decreasing | 0.46 (-1.63, 2.56) | | 0.54 (-1.34, 2.42) | | |
| | Average-high, decreasing | 1.89 (-0.17, 3.96) | | 0.60 (-1.24, 2.44) | | |
| | High, decreasing | -0.23 (-2.82, 2.35) | | -0.92 (-3.24, 1.40) | | |
| | High, decreasing fast | 1.12 (-1.59, 3.83) | | -0.27 (-2.80, 2.25) | | |
| AIx, % | $NO_2$ from pregnancy-18y | -0.42 (-1.30, 0.47) | 0.358 | 0.38 (-0.52, 1.27) | 0.407 | 0.321 |
| | Low, decreasing | Reference | 0.321 | Reference | 0.120 | 0.833 |
| | Average-low, decreasing | 1.80 (-0.78, 4.38) | | -0.38 (-3.01, 2.25) | | |
| | Average, decreasing | -0.45 (-2.89, 2.00) | | 1.54 (-0.93, 4.01) | | |
| | Average-high, decreasing | 0.44 (-1.97, 2.85) | | -0.40 (-2.82, 2.01) | | |
| | High, decreasing | -0.41 (-3.41, 2.59) | | 1.84 (-1.19, 4.88) | | |
| | High, decreasing fast | 1.29 (-1.87, 4.44) | | -1.12 (-4.43, 2.19) | | |

AIx: augmentation index; CIMT: carotid media-intima thickness; DBP: diastolic blood pressure; $NO_2$: nitrogen dioxide; PWV: pulse wave velocity; SBP: systolic blood pressure

**Supplementary Table 16.** Association between black carbon (BC) and cardiovascular health markers at 18 years in males and females.

| Outcome | Exposure | Males beta (95%CI) | p-value | Females beta (95%CI) | p-value | p-value sex interaction |
|---|---|---|---|---|---|---|
| Peripheral SBP, mmHg | BC from pregnancy-18y | 0.71 (-0.33, 1.75) | 0.183 | -0.38 (-1.23, 0.47) | 0.381 | 0.038 |
| | Low, decreasing | Reference | 0.334 | Reference | 0.076 | 0.239 |
| | Average-low, decreasing | 0.30 (-1.64, 2.24) | | 0.15 (-1.45, 1.75) | | |
| | Average, decreasing | 1.19 (-0.73, 3.11) | | -0.42 (-2.06, 1.22) | | |
| | Average-high, decreasing | 1.94 (-0.03, 3.90) | | 0.44 (-1.22, 2.09) | | |
| | High, decreasing | 1.24 (-1.05, 3.53) | | -0.71 (-2.56, 1.13) | | |
| | High, decreasing fast | 1.93 (-0.57, 4.43) | | 2.49 (0.30, 4.68) | | |
| Central SBP, mmHg | BC from pregnancy-18y | 0.69 (-0.15, 1.53) | 0.109 | -0.22 (-0.98, 0.53) | 0.564 | 0.034 |
| | Low, decreasing | Reference | 0.482 | Reference | 0.166 | 0.259 |
| | Average-low, decreasing | 0.40 (-1.18, 1.97) | | 0.57 (-0.85, 2.00) | | |
| | Average, decreasing | 1.03 (-0.53, 2.59) | | 0.23 (-1.23, 1.69) | | |
| | Average-high, decreasing | 1.45 (-0.14, 3.05) | | 0.79 (-0.68, 2.27) | | |
| | High, decreasing | 1.19 (-0.67, 3.05) | | -0.39 (-2.04, 1.25) | | |
| | High, decreasing fast | 1.36 (-0.67, 3.38) | | 2.10 (0.15, 4.05) | | |
| Peripheral DBP, mmHg | BC from pregnancy-18y | 0.78 (0.06, 1.51) | 0.035 | -0.07 (-0.71, 0.58) | 0.840 | 0.030 |
| | Low, decreasing | Reference | 0.164 | Reference | 0.127 | 0.127 |
| | Average-low, decreasing | 0.72 (-0.63, 2.08) | | 0.97 (-0.24, 2.18) | | |
| | Average, decreasing | 1.14 (-0.20, 2.48) | | 0.51 (-0.73, 1.76) | | |
| | Average-high, decreasing | 1.02 (-0.35, 2.39) | | 0.95 (-0.30, 2.21) | | |
| | High, decreasing | 2.21 (0.61, 3.80) | | -0.08 (-1.49, 1.32) | | |
| | High, decreasing fast | 1.31 (-0.43, 3.05) | | 1.86 (0.20, 3.52) | | |
| Central DBP, mmHg | BC from pregnancy-18y | 0.92 (0.18, 1.67) | 0.015 | -0.02 (-0.68, 0.64) | 0.955 | 0.024 |
| | Low, decreasing | Reference | 0.080 | Reference | 0.107 | 0.102 |
| | Average-low, decreasing | 0.80 (-0.59, 2.18) | | 1.01 (-0.23, 2.25) | | |
| | Average, decreasing | 1.34 (-0.04, 2.71) | | 0.55 (-0.73, 1.82) | | |
| | Average-high, decreasing | 1.18 (-0.23, 2.58) | | 1.12 (-0.17, 2.40) | | |
| | High, decreasing | 2.51 (0.87, 4.15) | | -0.06 (-1.49, 1.37) | | |

| | | | | | | |
|---|---|---|---|---|---|---|
| | High, decreasing fast | 1.50 (-0.28, 3.29) | | 1.89 (0.19, 3.59) | | |
| CIMT, µm | BC from pregnancy-18y | 1.87 (-2.20, 5.94) | 0.368 | 0.47 (-3.00, 3.94) | 0.790 | 0.717 |
| | Low, decreasing | Reference | 0.751 | Reference | 0.208 | 0.944 |
| | Average-low, decreasing | -0.72 (-8.18, 6.74) | | -5.51 (-11.98, 0.95) | | |
| | Average, decreasing | 1.42 (-6.18, 9.02) | | -5.44 (-12.03, 1.15) | | |
| | Average-high, decreasing | 3.66 (-4.00, 11.33) | | 0.45 (-6.23, 7.13) | | |
| | High, decreasing | 0.75 (-8.25, 9.75) | | -2.49 (-10.05, 5.07) | | |
| | High, decreasing fast | -3.14 (-13.22, 6.95) | | -5.08 (-13.76, 3.60) | | |
| PWV, m/s | BC from pregnancy-18y | -0.03 (-0.09, 0.04) | 0.460 | -0.02 (-0.07, 0.03) | 0.514 | 0.567 |
| | Low, decreasing | Reference | 0.083 | Reference | 0.056 | 0.532 |
| | Average-low, decreasing | 0.15 (0.03, 0.27) | | 0.13 (0.03, 0.23) | | |
| | Average, decreasing | 0.12 (0.00, 0.25) | | 0.01 (-0.09, 0.11) | | |
| | Average-high, decreasing | 0.04 (-0.09, 0.16) | | 0.02 (-0.08, 0.12) | | |
| | High, decreasing | 0.02 (-0.13, 0.17) | | 0.03 (-0.09, 0.15) | | |
| | High, decreasing fast | 0.00 (-0.16, 0.17) | | 0.00 (-0.13, 0.13) | | |
| Heart rate, bpm | BC from pregnancy-18y | 0.87 (-0.15, 1.88) | 0.095 | 0.49 (-0.41, 1.40) | 0.286 | 0.675 |
| | Low, decreasing | Reference | 0.048 | Reference | 0.315 | 0.690 |
| | Average-low, decreasing | 0.69 (-1.21, 2.59) | | 0.90 (-0.81, 2.61) | | |
| | Average, decreasing | 1.99 (0.11, 3.87) | | 0.51 (-1.24, 2.26) | | |
| | Average-high, decreasing | 0.48 (-1.44, 2.40) | | 1.70 (-0.07, 3.47) | | |
| | High, decreasing | 3.08 (0.84, 5.32) | | 0.72 (-1.25, 2.69) | | |
| | High, decreasing fast | 1.70 (-0.74, 4.15) | | 2.17 (-0.17, 4.51) | | |
| AIx, % | BC from pregnancy-18y | -0.66 (-1.84, 0.53) | 0.278 | -0.02 (-1.2, 1.17) | 0.976 | 0.702 |
| | Low, decreasing | Reference | 0.664 | Reference | 0.313 | 0.938 |
| | Average-low, decreasing | -0.27 (-2.47, 1.94) | | -0.78 (-3.01, 1.45) | | |
| | Average, decreasing | -1.41 (-3.59, 0.77) | | 0.64 (-1.65, 2.93) | | |
| | Average-high, decreasing | -1.17 (-3.40, 1.06) | | 0.53 (-1.78, 2.85) | | |
| | High, decreasing | -1.82 (-4.42, 0.78) | | -0.80 (-3.37, 1.77) | | |
| | High, decreasing fast | -1.11 (-3.96, 1.73) | | -2.16 (-5.20, 0.89) | | |

AIx: augmentation index; BC: black carbon; CIMT: carotid media-intima thickness; DBP: diastolic blood pressure; PWV: pulse wave velocity; SBP: systolic blood pressure

**Supplementary Table 17**. Association between fine particulate matter ($PM_{2.5}$) and cardiovascular health markers at 18 years in non-movers and movers.

| Outcome | Exposure | Non-movers beta (95%CI) | Non-movers p-value | Movers beta (95%CI) | Movers p-value | p-value interaction |
|---|---|---|---|---|---|---|
| Peripheral SBP, mmHg | $PM_{2.5}$ from pregnancy-18y | -0.10 (-0.66, 0.46) | 0.723 | 0.90 (-0.07, 1.88) | 0.070 | 0.191 |
| | Low, decreasing | Reference | 0.853 | Reference | 0.372 | 0.746 |
| | Average-low, decreasing | -0.27 (-2.82, 2.29) | | 0.21 (-2.02, 2.44) | | |
| | Average-high, decreasing | -0.69 (-3.07, 1.70) | | 0.89 (-1.55, 3.34) | | |
| | High, decreasing | -0.73 (-3.20, 1.74) | | 3.57 (-0.61, 7.75) | | |
| Central SBP, mmHg | $PM_{2.5}$ from pregnancy-18y | 0.08 (-0.39, 0.55) | 0.740 | 1.15 (0.30, 1.99) | 0.008 | 0.133 |
| | Low, decreasing | Reference | 0.934 | Reference | 0.178 | 0.499 |
| | Average-low, decreasing | 0.21 (-1.96, 2.37) | | 0.35 (-1.59, 2.28) | | |
| | Average-high, decreasing | -0.03 (-2.05, 2.00) | | 1.22 (-0.90, 3.34) | | |
| | High, decreasing | 0.21 (-1.88, 2.30) | | 3.80 (0.18, 7.43) | | |
| Peripheral DBP, mmHg | $PM_{2.5}$ from pregnancy-18y | 0.30 (-0.10, 0.70) | 0.144 | 1.22 (0.47, 1.96) | 0.002 | 0.194 |
| | Low, decreasing | Reference | 0.699 | Reference | 0.093 | 0.328 |
| | Average-low, decreasing | 0.40 (-1.43, 2.24) | | 0.53 (-1.17, 2.24) | | |
| | Average-high, decreasing | 0.41 (-1.30, 2.12) | | 0.91 (-0.96, 2.77) | | |
| | High, decreasing | 0.77 (-1.00, 2.55) | | 4.09 (0.90, 7.28) | | |
| Central DBP, mmHg | $PM_{2.5}$ from pregnancy-18y | 0.35 (-0.06, 0.76) | 0.098 | 1.23 (0.46, 2.00) | 0.002 | 0.222 |
| | Low, decreasing | Reference | 0.645 | Reference | 0.102 | 0.381 |
| | Average-low, decreasing | 0.53 (-1.35, 2.41) | | 0.59 (-1.17, 2.34) | | |
| | Average-high, decreasing | 0.62 (-1.13, 2.38) | | 0.85 (-1.07, 2.77) | | |
| | High, decreasing | 0.96 (-0.85, 2.78) | | 4.15 (0.87, 7.43) | | |
| CIMT, µm | $PM_{2.5}$ from pregnancy-18y | -1.82 (-4.03, 0.38) | 0.106 | -5.76 (-9.77, -1.76) | 0.005 | 0.635 |
| | Low, decreasing | Reference | 0.477 | Reference | 0.015 | 0.096 |
| | Average-low, decreasing | 2.15 (-7.79, 12.09) | | -8.29 (-17.36, 0.79) | | |
| | Average-high, decreasing | -0.20 (-9.46, 9.07) | | -11.63 (-21.38, -1.87) | | |
| | High, decreasing | -2.17 (-11.76, 7.42) | | -24.75 (-41.88, -7.61) | | |
| PWV, m/s | $PM_{2.5}$ from pregnancy-18y | 0.01 (-0.03, 0.04) | 0.671 | 0.05 (-0.01, 0.11) | 0.096 | 0.279 |
| | Low, decreasing | Reference | 0.527 | Reference | 0.253 | 0.430 |

| | | | | | | |
|---|---|---|---|---|---|---|
| | Average-low, decreasing | 0.03 (-0.13, 0.20) | | 0.04 (-0.09, 0.18) | | |
| | Average-high, decreasing | 0.07 (-0.08, 0.23) | | 0.01 (-0.13, 0.15) | | |
| | High, decreasing | 0.04 (-0.12, 0.20) | | 0.23 (-0.01, 0.47) | | |
| Heart rate, bpm | $PM_{2.5}$ from pregnancy-18y | 0.26 (-0.31, 0.82) | 0.374 | 0.72 (-0.33, 1.76) | 0.177 | 0.785 |
| | Low, decreasing | Reference | 0.250 | Reference | 0.443 | 0.808 |
| | Average-low, decreasing | -1.15 (-3.76, 1.45) | | 1.87 (-0.46, 4.20) | | |
| | Average-high, decreasing | 0.05 (-2.38, 2.48) | | 0.79 (-1.76, 3.34) | | |
| | High, decreasing | -0.48 (-3.00, 2.04) | | 1.72 (-2.65, 6.09) | | |
| AIx, % | $PM_{2.5}$ from pregnancy-18y | -0.17 (-0.88, 0.55) | 0.651 | 0.28 (-0.91, 1.47) | 0.645 | 0.634 |
| | Low, decreasing | Reference | 0.408 | Reference | 0.590 | 0.694 |
| | Average-low, decreasing | 1.24 (-2.05, 4.53) | | -1.25 (-3.95, 1.44) | | |
| | Average-high, decreasing | 0.37 (-2.70, 3.44) | | 0.54 (-2.43, 3.51) | | |
| | High, decreasing | 1.28 (-1.90, 4.46) | | -0.84 (-5.86, 4.18) | | |

AIx: augmentation index; CIMT: carotid media-intima thickness; DBP: diastolic blood pressure; $PM_{2.5}$: fine particulate matter; PWV: pulse wave velocity; SBP: systolic blood pressure

**Supplementary Table 18.** Association between nitrogen dioxide ($NO_2$) and cardiovascular health markers at 18 years in non-movers and movers.

| Outcome | Exposure | Non-movers beta (95%CI) | Non-movers p-value | Movers beta (95%CI) | Movers p-value | p-value interaction |
|---|---|---|---|---|---|---|
| Peripheral SBP, mmHg | $NO_2$ from pregnancy-18y | -0.02 (-0.59, 0.56) | 0.957 | 0.76 (-0.45, 1.97) | 0.220 | 0.401 |
| | Low, decreasing | Reference | 0.336 | Reference | 0.222 | 0.290 |
| | Average-low, decreasing | -0.44 (-2.18, 1.30) | | -0.75 (-3.79, 2.29) | | |
| | Average, decreasing | 0.28 (-1.36, 1.92) | | -0.21 (-3.40, 2.97) | | |
| | Average-high, decreasing | -0.34 (-1.95, 1.27) | | -2.37 (-6.00, 1.27) | | |
| | High, decreasing | 0.26 (-1.67, 2.19) | | 4.16 (-1.24, 9.56) | | |
| | High, decreasing fast | 2.15 (-0.58, 4.87) | | 1.16 (-1.63, 3.96) | | |
| Central SBP, mmHg | $NO_2$ from pregnancy-18y | 0.13 (-0.36, 0.62) | 0.591 | 1.01 (-0.04, 2.06) | 0.058 | 0.295 |
| | Low, decreasing | Reference | 0.141 | Reference | 0.306 | 0.193 |
| | Average-low, decreasing | 0.18 (-1.29, 1.66) | | -0.12 (-2.63, 2.39) | | |
| | Average, decreasing | 0.53 (-0.86, 1.93) | | 1.16 (-1.47, 3.80) | | |
| | Average-high, decreasing | 0.26 (-1.11, 1.63) | | -1.49 (-4.49, 1.52) | | |
| | High, decreasing | 1.01 (-0.63, 2.65) | | 3.52 (-0.95, 7.98) | | |
| | High, decreasing fast | 2.88 (0.57, 5.19) | | 0.93 (-1.38, 3.24) | | |
| Peripheral DBP, mmHg | $NO_2$ from pregnancy-18y | 0.30 (-0.11, 0.72) | 0.155 | 1.20 (0.27, 2.13) | 0.012 | 0.224 |
| | Low, decreasing | Reference | 0.077 | Reference | 0.244 | 0.409 |
| | Average-low, decreasing | 0.73 (-0.52, 1.99) | | 0.99 (-1.15, 3.12) | | |
| | Average, decreasing | 0.57 (-0.61, 1.75) | | 2.55 (0.31, 4.79) | | |
| | Average-high, decreasing | 0.73 (-0.43, 1.89) | | 0.30 (-2.25, 2.85) | | |
| | High, decreasing | 1.50 (0.11, 2.89) | | 2.95 (-0.84, 6.75) | | |
| | High, decreasing fast | 2.64 (0.68, 4.61) | | 1.05 (-0.91, 3.01) | | |
| Central DBP, mmHg | $NO_2$ from pregnancy-18y | 0.36 (-0.07, 0.78) | 0.101 | 1.19 (0.23, 2.15) | 0.015 | 0.254 |
| | Low, decreasing | Reference | 0.045 | Reference | 0.343 | 0.320 |
| | Average-low, decreasing | 0.79 (-0.49, 2.08) | | 0.90 (-1.30, 3.10) | | |
| | Average, decreasing | 0.71 (-0.50, 1.92) | | 2.49 (0.18, 4.79) | | |
| | Average-high, decreasing | 0.94 (-0.25, 2.13) | | 0.43 (-2.20, 3.05) | | |
| | High, decreasing | 1.62 (0.20, 3.04) | | 2.65 (-1.25, 6.55) | | |

| | | | | | | |
|---|---|---|---|---|---|---|
| | High, decreasing fast | 3.00 (1.00, 5.01) | | 0.98 (-1.04, 2.99) | | |
| CIMT, µm | $NO_2$ from pregnancy-18y | -0.05 (-2.35, 2.24) | 0.964 | -2.30 (-7.31, 2.72) | 0.369 | 0.640 |
| | Low, decreasing | Reference | 0.768 | Reference | 0.648 | 0.819 |
| | Average-low, decreasing | -2.34 (-9.22, 4.54) | | 1.38 (-10.36, 13.11) | | |
| | Average, decreasing | -1.42 (-7.94, 5.10) | | -4.61 (-16.72, 7.51) | | |
| | Average-high, decreasing | -0.32 (-6.73, 6.10) | | -8.01 (-22.17, 6.15) | | |
| | High, decreasing | -4.12 (-11.82, 3.57) | | -1.49 (-21.71, 18.72) | | |
| | High, decreasing fast | -4.17 (-15.19, 6.84) | | 2.59 (-8.21, 13.40) | | |
| PWV, m/s | $NO_2$ from pregnancy-18y | 0.00 (-0.04, 0.03) | 0.846 | 0.02 (-0.05, 0.09) | 0.613 | 0.449 |
| | Low, decreasing | Reference | 0.284 | Reference | 0.479 | 0.096 |
| | Average-low, decreasing | 0.09 (-0.02, 0.20) | | 0.10 (-0.08, 0.27) | | |
| | Average, decreasing | 0.11 (0.01, 0.21) | | -0.06 (-0.24, 0.12) | | |
| | Average-high, decreasing | 0.07 (-0.03, 0.18) | | -0.09 (-0.30, 0.12) | | |
| | High, decreasing | 0.04 (-0.09, 0.16) | | 0.02 (-0.28, 0.32) | | |
| | High, decreasing fast | 0.12 (-0.05, 0.29) | | -0.04 (-0.20, 0.13) | | |
| Heart rate, bpm | $NO_2$ from pregnancy-18y | 0.01 (-0.58, 0.60) | 0.973 | 0.98 (-0.31, 2.27) | 0.137 | 0.354 |
| | Low, decreasing | Reference | 0.309 | Reference | 0.084 | 0.960 |
| | Average-low, decreasing | 0.44 (-1.34, 2.21) | | 0.93 (-2.12, 3.98) | | |
| | Average, decreasing | -0.08 (-1.75, 1.60) | | 3.66 (0.47, 6.85) | | |
| | Average-high, decreasing | 0.84 (-0.8, 2.49) | | 3.00 (-0.65, 6.64) | | |
| | High, decreasing | -0.70 (-2.67, 1.27) | | -2.66 (-8.07, 2.75) | | |
| | High, decreasing fast | 0.74 (-2.03, 3.52) | | 0.31 (-2.49, 3.11) | | |
| AIx, % | $NO_2$ from pregnancy-18y | 0.08 (-0.66, 0.82) | 0.834 | -0.1 (-1.57, 1.36) | 0.890 | 0.771 |
| | Low, decreasing | Reference | 0.663 | Reference | 0.881 | 0.955 |
| | Average-low, decreasing | 1.27 (-0.97, 3.51) | | -0.58 (-3.98, 2.83) | | |
| | Average, decreasing | 1.40 (-0.72, 3.51) | | -0.50 (-4.04, 3.04) | | |
| | Average-high, decreasing | 0.64 (-1.44, 2.72) | | -0.81 (-4.86, 3.24) | | |
| | High, decreasing | 1.70 (-0.78, 4.19) | | -2.77 (-8.73, 3.19) | | |
| | High, decreasing fast | 1.22 (-2.29, 4.74) | | -1.75 (-4.86, 1.36) | | |

AIx: augmentation index; CIMT: carotid media-intima thickness; DBP: diastolic blood pressure; $NO_2$: nitrogen dioxide; PWV: pulse wave velocity; SBP: systolic blood pressure

**Supplementary Table 19.** Association between black carbon (BC) and cardiovascular health markers at 18 years in non-movers and movers.

| Outcome | Exposure | Non-movers beta (95%CI) | p-value | Movers beta (95%CI) | p-value | p-value interaction |
|---|---|---|---|---|---|---|
| Peripheral SBP, mmHg | BC from pregnancy-18y | -0.09 (-0.84, 0.66) | 0.816 | 0.81 (-1.11, 2.74) | 0.406 | 0.523 |
| | Low, decreasing | Reference | 0.071 | Reference | 0.703 | 0.237 |
| | Average-low, decreasing | 0.30 (-1.20, 1.79) | | -0.22 (-2.72, 2.27) | | |
| | Average, decreasing | 0.14 (-1.35, 1.63) | | 0.76 (-2.48, 4.00) | | |
| | Average-high, decreasing | 1.04 (-0.47, 2.55) | | 0.58 (-2.92, 4.08) | | |
| | High, decreasing | -0.07 (-1.73, 1.60) | | 1.55 (-4.40, 7.50) | | |
| | High, decreasing fast | 2.83 (0.57, 5.09) | | 1.81 (-0.58, 4.21) | | |
| Central SBP, mmHg | BC from pregnancy-18y | -0.01 (-0.64, 0.62) | 0.974 | 1.00 (-0.67, 2.67) | 0.241 | 0.407 |
| | Low, decreasing | Reference | 0.122 | Reference | 0.886 | 0.264 |
| | Average-low, decreasing | 0.50 (-0.77, 1.77) | | 0.48 (-1.69, 2.66) | | |
| | Average, decreasing | 0.51 (-0.76, 1.77) | | 0.54 (-2.29, 3.36) | | |
| | Average-high, decreasing | 0.99 (-0.29, 2.27) | | 1.37 (-1.68, 4.42) | | |
| | High, decreasing | 0.13 (-1.29, 1.54) | | 1.11 (-4.07, 6.30) | | |
| | High, decreasing fast | 2.42 (0.50, 4.34) | | 1.24 (-0.85, 3.33) | | |
| Peripheral DBP, mmHg | BC from pregnancy-18y | 0.19 (-0.35, 0.73) | 0.482 | 0.98 (-0.50, 2.46) | 0.195 | 0.430 |
| | Low, decreasing | Reference | 0.512 | Reference | 0.335 | 0.704 |
| | Average-low, decreasing | 0.62 (-0.46, 1.70) | | 1.84 (-0.09, 3.77) | | |
| | Average, decreasing | 0.69 (-0.39, 1.76) | | 0.29 (-2.21, 2.79) | | |
| | Average-high, decreasing | 0.80 (-0.29, 1.89) | | 1.28 (-1.42, 3.98) | | |
| | High, decreasing | 0.65 (-0.55, 1.86) | | 1.99 (-2.60, 6.58) | | |
| | High, decreasing fast | 1.64 (0.01, 3.27) | | 1.77 (-0.07, 3.62) | | |
| Central DBP, mmHg | BC from pregnancy-18y | 0.29 (-0.26, 0.84) | 0.304 | 0.99 (-0.53, 2.52) | 0.200 | 0.468 |
| | Low, decreasing | Reference | 0.346 | Reference | 0.365 | 0.606 |
| | Average-low, decreasing | 0.72 (-0.39, 1.82) | | 1.85 (-0.14, 3.83) | | |
| | Average, decreasing | 0.85 (-0.25, 1.95) | | 0.14 (-2.43, 2.71) | | |
| | Average-high, decreasing | 1.00 (-0.11, 2.12) | | 1.44 (-1.33, 4.22) | | |
| | High, decreasing | 0.84 (-0.39, 2.08) | | 1.90 (-2.82, 6.62) | | |

| | | | | | | |
|---|---|---|---|---|---|---|
| | High, decreasing fast | 1.88 (0.21, 3.55) | | 1.71 (-0.19, 3.61) | | |
| CIMT, µm | BC from pregnancy-18y | 1.72 (-1.24, 4.68) | 0.255 | -2.48 (-10.36, 5.41) | 0.537 | 0.465 |
| | Low, decreasing | Reference | 0.229 | Reference | 0.663 | 0.917 |
| | Average-low, decreasing | -2.59 (-8.44, 3.26) | | -5.39 (-15.55, 4.77) | | |
| | Average, decreasing | -2.32 (-8.18, 3.55) | | 3.53 (-10.09, 17.15) | | |
| | Average-high, decreasing | 2.85 (-3.07, 8.78) | | -3.06 (-17.34, 11.22) | | |
| | High, decreasing | -0.22 (-6.84, 6.41) | | 0.73 (-22.64, 24.10) | | |
| | High, decreasing fast | -2.88 (-11.94, 6.18) | | -6.45 (-16.30, 3.41) | | |
| PWV, m/s | BC from pregnancy-18y | -0.04 (-0.09, 0.01) | 0.111 | -0.01 (-0.13, 0.10) | 0.851 | 0.402 |
| | Low, decreasing | Reference | 0.016 | Reference | 0.229 | 0.051 |
| | Average-low, decreasing | 0.15 (0.05, 0.24) | | 0.11 (-0.03, 0.26) | | |
| | Average, decreasing | 0.07 (-0.02, 0.17) | | -0.02 (-0.22, 0.17) | | |
| | Average-high, decreasing | 0.04 (-0.05, 0.14) | | -0.09 (-0.30, 0.12) | | |
| | High, decreasing | 0.03 (-0.08, 0.13) | | 0.12 (-0.23, 0.47) | | |
| | High, decreasing fast | 0.08 (-0.06, 0.22) | | -0.07 (-0.22, 0.07) | | |
| Heart rate, bpm | BC from pregnancy-18y | 0.66 (-0.10, 1.42) | 0.087 | 1.15 (-0.90, 3.19) | 0.270 | 0.891 |
| | Low, decreasing | Reference | 0.363 | Reference | 0.541 | 0.973 |
| | Average-low, decreasing | 0.72 (-0.80, 2.24) | | 1.42 (-1.24, 4.08) | | |
| | Average, decreasing | 1.21 (-0.31, 2.73) | | 0.32 (-3.13, 3.77) | | |
| | Average-high, decreasing | 1.08 (-0.46, 2.62) | | 2.57 (-1.15, 6.29) | | |
| | High, decreasing | 1.76 (0.06, 3.46) | | -0.40 (-6.73, 5.93) | | |
| | High, decreasing fast | 1.94 (-0.37, 4.25) | | 2.09 (-0.46, 4.64) | | |
| AIx, % | BC from pregnancy-18y | -0.26 (-1.22, 0.70) | 0.600 | -0.56 (-2.88, 1.75) | 0.632 | 0.849 |
| | Low, decreasing | Reference | 0.965 | Reference | 0.063 | 0.966 |
| | Average-low, decreasing | -0.28 (-2.20, 1.65) | | -1.10 (-4.11, 1.91) | | |
| | Average, decreasing | 0.02 (-1.90, 1.94) | | -0.33 (-4.19, 3.52) | | |
| | Average-high, decreasing | -0.11 (-2.05, 1.83) | | 2.47 (-1.74, 6.68) | | |
| | High, decreasing | -0.76 (-2.90, 1.38) | | -6.59 (-13.66, 0.49) | | |
| | High, decreasing fast | -0.55 (-3.46, 2.36) | | -3.03 (-5.89, -0.16) | | |

AIx: augmentation index; BC: black carbon; CIMT: carotid media-intima thickness; DBP: diastolic blood pressure; PWV: pulse wave velocity; SBP: systolic blood pressure

**Supplementary Table 20.** Association between air pollutants from pregnancy to 18 years with birthweight and maternal pre-pregnancy body mass index (BMI).

| Exposure | | Birthweight | | Maternal BMI | |
|---|---|---|---|---|---|
| | | β (95% CI) | p-value | β (95% CI) | p-value |
| $PM_{2.5}$ | $PM_{2.5}$ from pregnancy-18y | -0.01 (-0.03, 0.01) | 0.577 | 0.06 (-0.08, 0.21) | 0.400 |
| | Low, decreasing | Reference | 0.470 | Reference | 0.607 |
| | Average-low, decreasing | -0.06 (-0.13, 0.02) | | 0.16 (-0.37, 0.70) | |
| | Average-high, decreasing | -0.04 (-0.11, 0.03) | | 0.28 (-0.20, 0.77) | |
| | High, decreasing | -0.05 (-0.12, 0.03) | | 0.15 (-0.37, 0.68) | |
| $NO_2$ | $NO_2$ from pregnancy-18y | -0.01 (-0.03, 0.01) | 0.391 | -0.01 (-0.17, 0.16) | 0.943 |
| | Low, decreasing | Reference | 0.752 | Reference | 0.240 |
| | Average-low, decreasing | 0.02 (-0.05, 0.08) | | 0.20 (-0.29, 0.68) | |
| | Average, decreasing | -0.02 (-0.08, 0.05) | | -0.06 (-0.52, 0.39) | |
| | Average-high, decreasing | 0.00 (-0.06, 0.06) | | 0.03 (-0.41, 0.48) | |
| | High, decreasing | -0.03 (-0.11, 0.05) | | -0.13 (-0.69, 0.43) | |
| | High, decreasing fast | -0.02 (-0.10, 0.07) | | -0.50 (-1.10, 0.10) | |
| BC | BC from pregnancy-18y | -0.01 (-0.04, 0.02) | 0.678 | 0.04 (-0.18, 0.26) | 0.696 |
| | Low, decreasing | Reference | 0.714 | Reference | 0.148 |
| | Average-low, decreasing | 0.02 (-0.04, 0.07) | | 0.17 (-0.24, 0.57) | |
| | Average, decreasing | -0.02 (-0.08, 0.03) | | -0.12 (-0.54, 0.30) | |
| | Average-high, decreasing | 0.00 (-0.06, 0.05) | | -0.15 (-0.57, 0.27) | |
| | High, decreasing | 0.00 (-0.07, 0.07) | | 0.24 (-0.24, 0.72) | |
| | High, decreasing fast | 0.02 (-0.05, 0.10) | | -0.36 (-0.91, 0.18) | |

BC: black carbon; BMI: body mass index; $NO_2$: nitrogen dioxide; $PM_{2.5}$: fine particulate matter

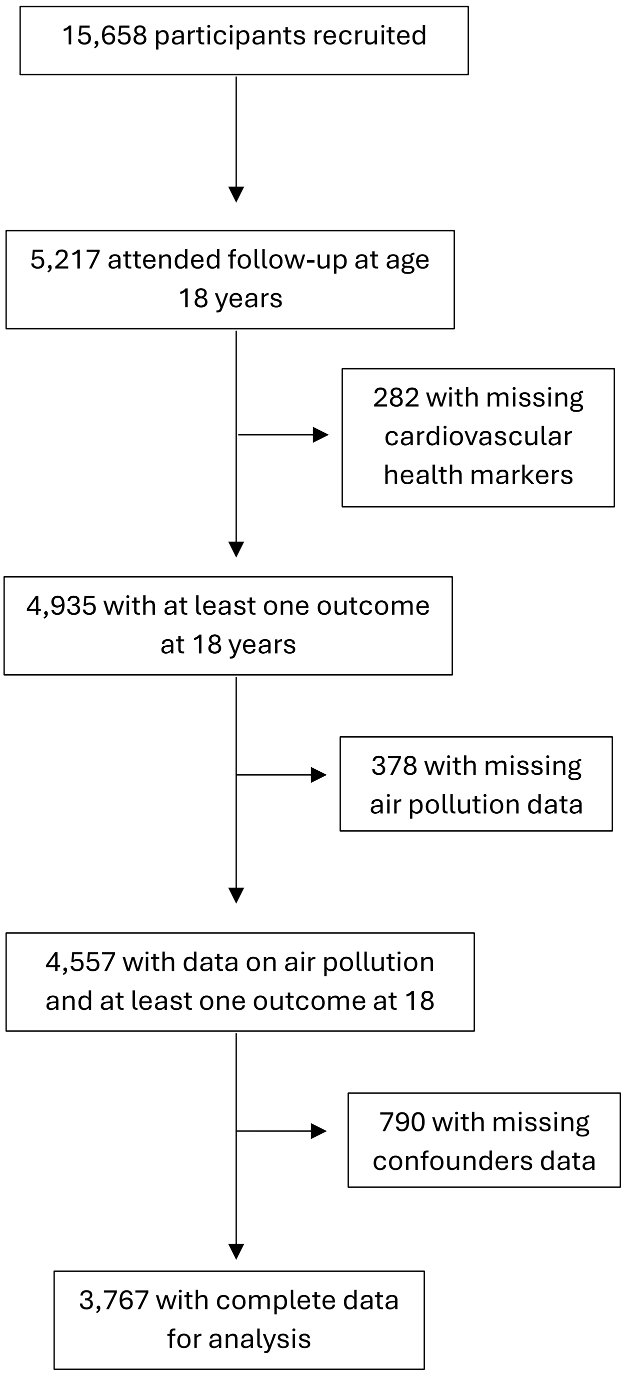


**Supplementary Figure 1.** Flow chart of participants.

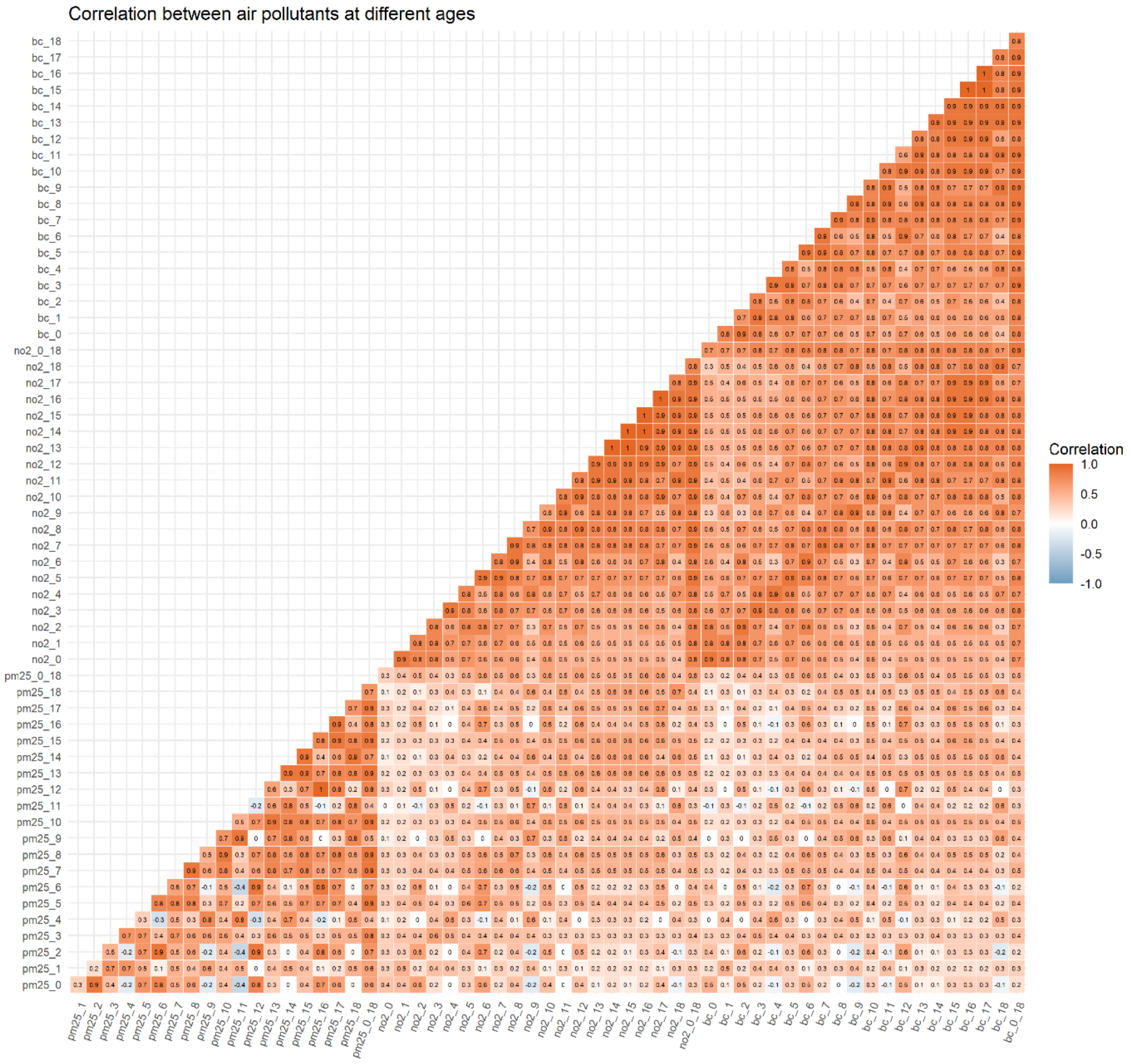


**Supplementary Figure 2.** Correlation between air pollutants across ages from pregnancy (age 0) to 18 years.

bc: black carbon; no2: nitrogen dioxide; pm25: fine particulate matter

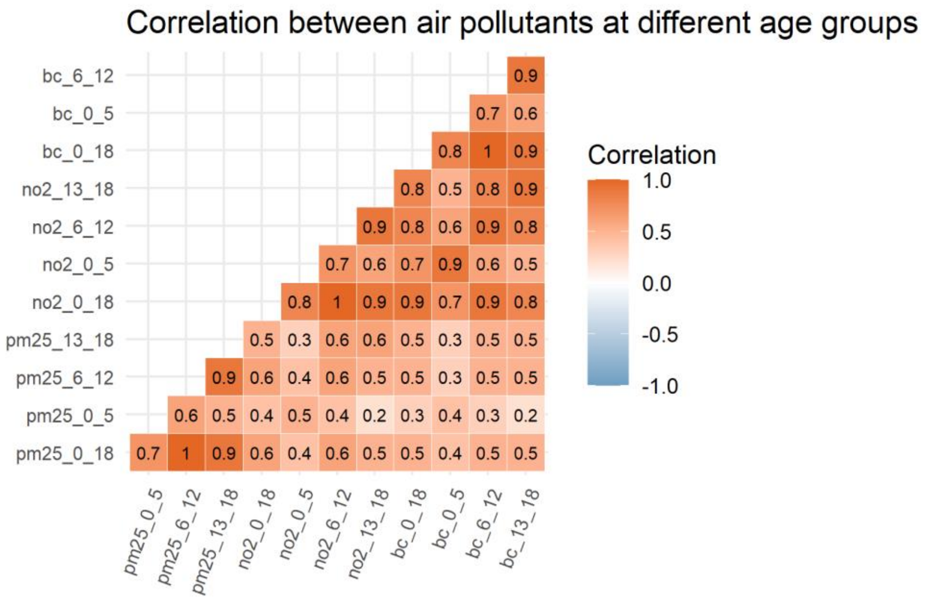


**Supplementary Figure 3.** Correlation between air pollutants across age ranges.

bc: black carbon; no2: nitrogen dioxide; pm25: fine particulate matter

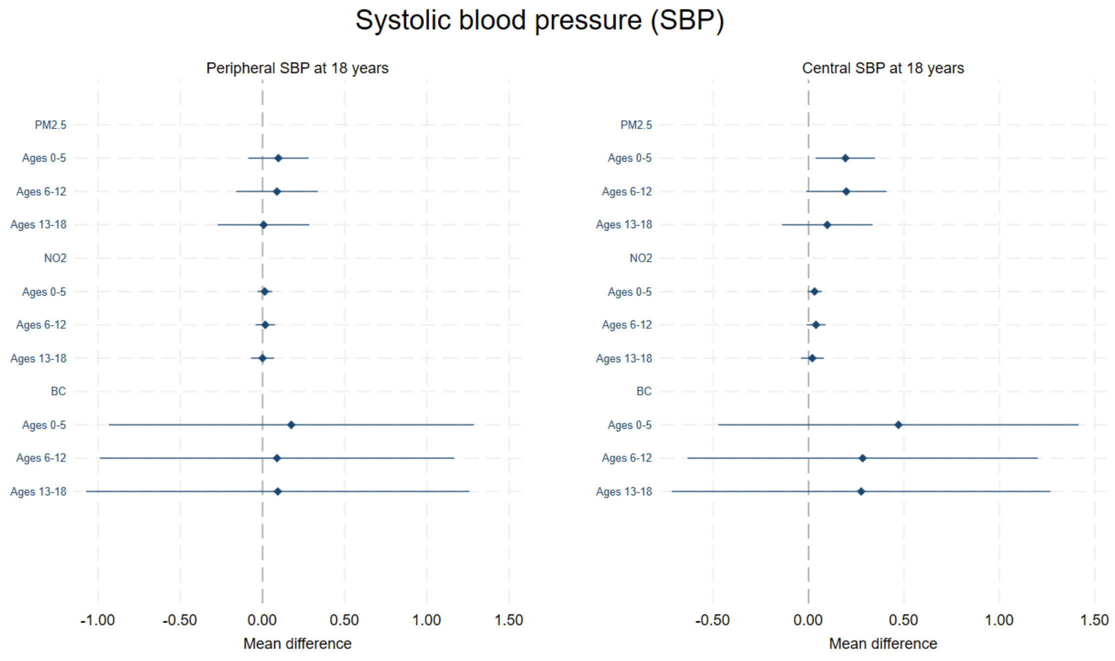


Adjusted for area deprivation, maternal education, maternal age, maternal ethnicity, age, and sex

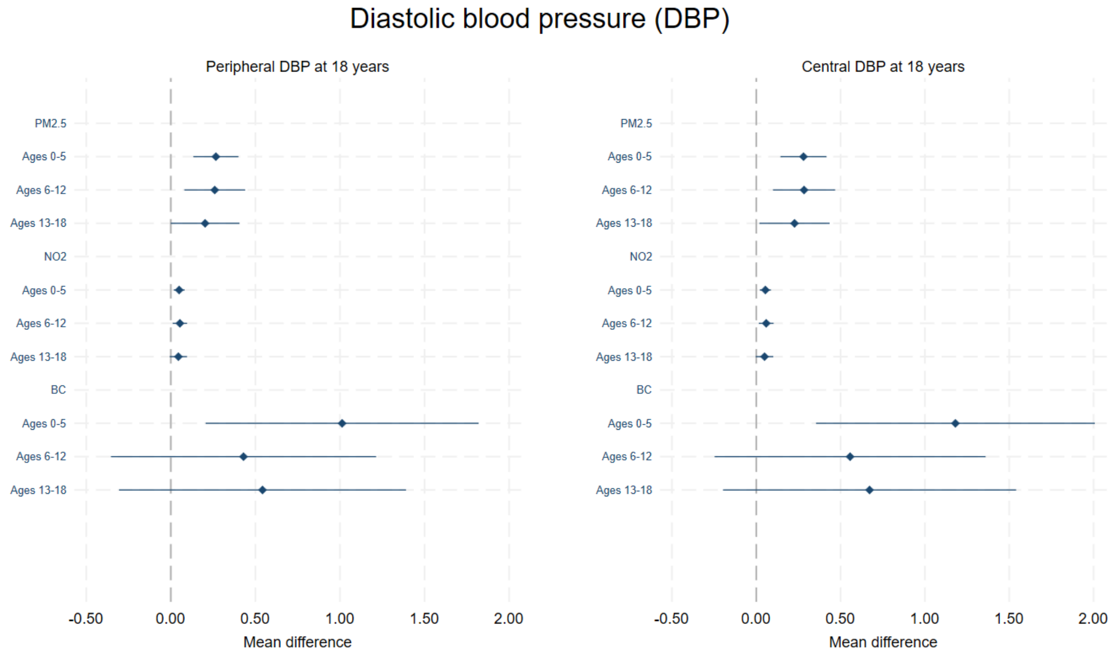


Adjusted for area deprivation, maternal education, maternal age, maternal ethnicity, age, and sex

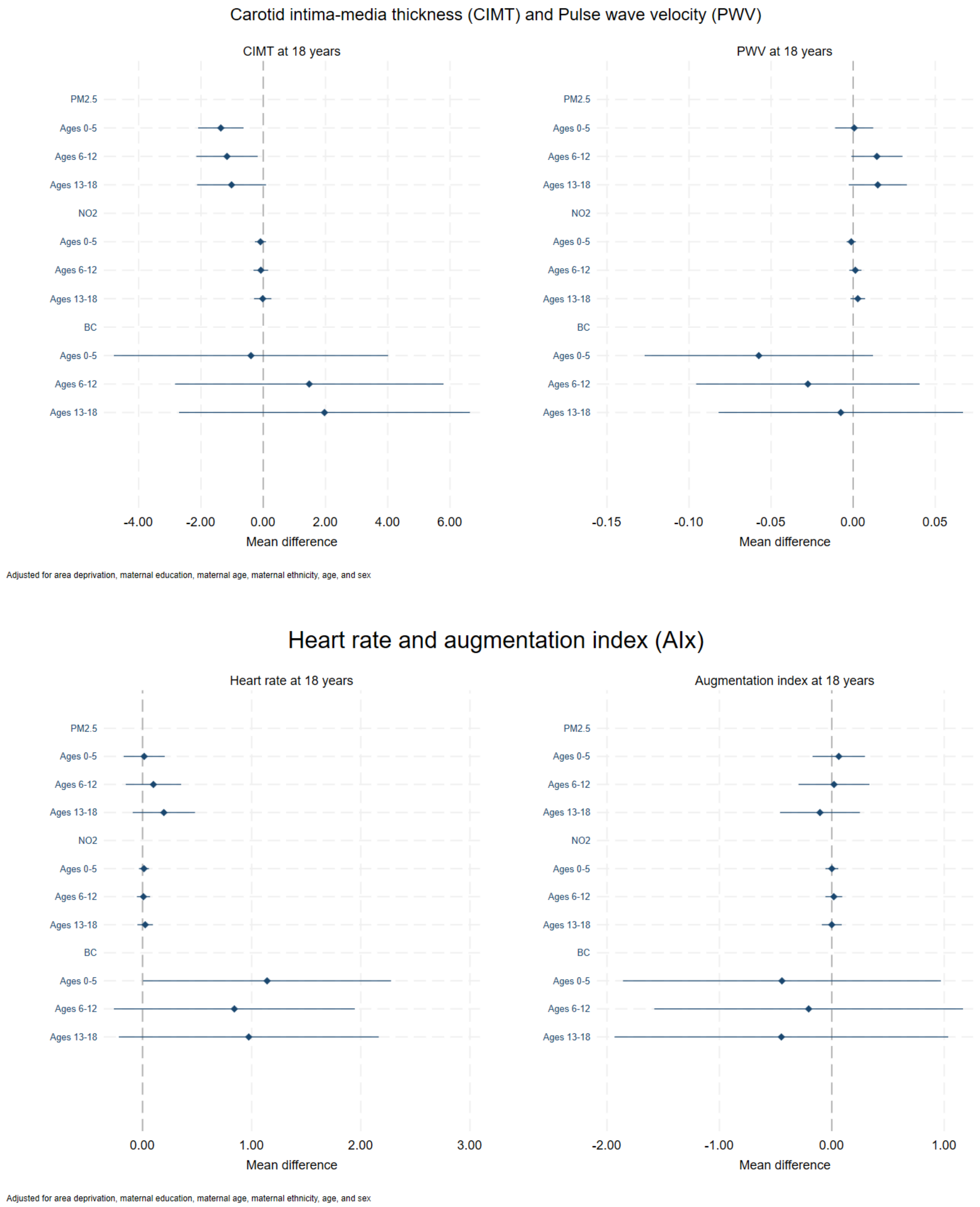


**Supplementary Figure 4.** Association between air pollution at different age ranges and cardiovascular health markers at age 18 years.

Estimates are mean difference in the outcome per unit increase in the exposure in each age period.
BC: black carbon; BMI: body mass index; $NO_2$: nitrogen dioxide; $PM_{2.5}$: fine particulate matter
